# Targeting OAS3 for reversing M2d infiltration and restoring anti-tumor immunity in pancreatic cancer

**DOI:** 10.1101/2024.08.07.24311609

**Authors:** Shaopeng Zhang, Ximo Xu, Kundong Zhang, Changzheng Lei, Yitian Xu, Pengshan Zhang, Yuan Zhang, Haitao Gu, Chen Huang, Zhengjun Qiu

**Affiliations:** Department of Gastrointestinal Surgery, Shanghai General Hospital, Shanghai Jiaotong University School of Medicine, Shanghai, China; Department of General Surgery, Jiuquan Branch of Shanghai General Hospital, Gansu, China

**Keywords:** Pancreatic cancer, M2d, OAS3, Lactate, METTL3

## Abstract

Abundant infiltration of tumor-associated macrophages (TAMs) within the tumor stroma plays a pivotal role in inducing immune escape in pancreatic cancer (PC). Lactate serves as a direct regulator of macrophage polarization and functions, although the precise regulation mechanism remain inadequately understood. Our study revealed that PC cells promote macrophage polarization towards the M2d phenotype through high lactate secretion. M2d is characterized by elevated secretion of IL-10 and VEGF-A, which diminish CD8^+^ T cells cytotoxicity and promote tumor neoangiogenesis simultaneously. Additionally, we identify 2,5’-oligoadenylate synthase 3 (OAS3) as an essential regulator of M2d polarization, upregulated by PC cells via lactate/METTL3/OAS3 axis. METTL3 mediated m^6^A modification on OAS3 mRNA correlates with increased OAS3 expression in TAMs, which is associated with poorer prognosis in PC patients. OAS3 deficiency in macrophages substantially impairs IL-10^high^VEGF-A^high^M2d polarization and their pro-tumor functions while enhancing the therapeutic efficacy of gemcitabine (Gem) and anti-PD-L1 mAb in humanized mouse models. In conclusion, OAS3 presents as a promising immune therapeutic target for reversing IL-10^high^VEGF-A^high^M2d infiltration and restoring CD8^+^ T cell mediated anti-tumor immunity in pancreatic cancer.

## Introduction

Pancreatic cancer (PC) still carries the worst survival rate among all solid tumours, mostly due to the absence of early symptoms and high resistance to chemo- and radiotherapy, characterized by an alarmingly low 5-year survival rate of just 11%(1). A key characteristic of pancreatic tumors is their dense stromal composition, which significantly impedes the efficacy of immune checkpoint inhibitors (ICBs), otherwise successful in various cancers(2). The stromal components of PC predominantly consist of cancer-associated fibroblasts (CAFs), tumor-associated macrophages (TAMs), regulatory T cells (Tregs) and neutrophils, all of which contribute significantly to the aggressiveness and treatment resistance of PC(3,4).

Macrophages exhibit high plasticity and can polarize into two extreme polarization phenotypes: the “classically” activated macrophage (M1) and “alternatively” activated macrophage (M2). M1 cells have anti-tumour behaviors, while M2 cells can promote tumour growth, angiogenesis, invasion and metastasis(5). It was proposed that once the tumor is initiated and progresses toward malignancy, the macrophage phenotype shifts from M1 to M2(6). The majority of TAMs in PC patients were described to share phenotypical and functional features with CD163^+^CD206^+^M2 cells, increased presence of M2-like TAMs in PC is associated with increased metastasis and poor outcome(7). TAMs are recognized as dominant populations in tumors which suppress T cell function by releasing various oncogenic signals, including growth factors, proteases, and cytokines. TAMs derived IL-10 inhibits dendritic cell maturation and reduces antigen presentation, thereby suppressing anti-tumor immune responses and facilitating immune escape(8). Under the stimulation of pancreatic stellate cells (PSCs), TAMs can produce a large amount of cytokines that activate α-smooth muscle actin (α-SMA)-expressing myofibroblasts, promoting the formation of fibrosis in PC tumor microenvironment (TME)(9). The TGF-β secreted by TAMs can upregulate the phosphorylation of PKM2 in PC cells and co-activate STAT1, ultimately leading to the upregulation of PD-L1(10). Immunosuppressive cytokines derived from TAMs are major factors in hindering the function of cytotoxic T lymphocytes, fostering a profoundly immune-suppressed tumor environment and diminishes responsiveness to ICB therapy(11).

However, various immunotherapeutic methods targeting M2 have not yet achieved satisfactory results in PC treatment. Thus, gaining a deeper understanding of the TAMs within PC are essential for reversing the immunosuppressive TME and enhancing ICB therapy. Based on the differences in surface markers and cytokine secretion, M2 have been further subdivided into M2a, M2b, M2c and M2d. Both M2c and M2d have the ability to suppress immune responses and stimulate tumor growth. M2d promote angiogenesis and immune suppression by secreting a variety of proteases (such as matrix metalloproteinase-2), cytokines (such as VEGF), and anti-inflammatory factors (such as TGF-β and IL-10)(12). Hypoxia and high lactate are two major characteristic features of PC, Carlos. found that hypoxia increased VEGF-A secretion, an effect amplified by lactate, indicating that hypoxia and lactate regulate macrophage cytokine production in a synergistic manner(13). Lactate is the end product of aerobic glycolysis in tumor cells (Warburg effect). Oscar R. and colleagues first discovered that lactate can mediate M2 polarization, and this occurs not through the classical pathway dependent on IL-4 and IL-13. Lactate in the supernatant of tumor cell cultures can promote M2 polarization and the expression of M2 markers such as VEGF and Arg1 by regulating HIF-1α(14). While most studies have focused on lactate’s role in promoting M2 polarization, the specific mechanisms of M2 infiltration in PC tissues remain largely unknown(15).

N6-methyladenosine (m^6^A), as an mRNA modification, is abundant in nearly all eukaryotes, METTL3, serving as a catalytic subunit of the methyltransferase complex, has been reported to participate in diverse biological processes(16). lactylation was reported to drive oncogenesis by facilitating METTL3 in colon cancer-infiltrating myeloid cells, which is critical for the polarization of CD206^+^ M2-like TAMs by activating JAK1/STAT3 axis in TAMs(17). In contrast to the conclusion above, Huilong reported that METTL3-deficient mice show increased M1/M2-like TAMs and Tregs infiltration into tumours which enhances the activation of NF-kB and STAT3 through the ERK pathway, leading to increased tumour growth and metastasis(18). 2,5’-oligoadenylate synthase (OAS) is a class of enzymes induced by interferons and mainly encoded by the OAS1, OAS2, and OAS3 genes. OAS3 expression is associated with the prognosis and chemotherapeutic outcomes of various cancers, in addition, high OAS3 expression was proved to be related to worse overall survival time of PC patients(19,20). In terms of immune-infiltrating levels, OAS3 expression is positively associated with various infiltration of immunosuppressive cells including neutrophils, Tregs, CAFs and TAMs(19). However, the effect of OAS3 on tumor-infiltrating macrophages in the setting of high lactate TME has not been elucidated.

In this study, we discovered that PC cells polarize macrophages toward the tumor-promoting M2d phenotype *in vitro*. This polarization is characterized by increased expression of IL-10 and VEGF-A, which separately inhibit the tumor-killing ability of CD8^+^ T cells and promote neoangiogenesis. Furthermore, we revealed that lactate within the PC TME could be transferred into tumor-infiltrating macrophages to induce M2d polarization by upregulating METTL3, which is critical for capturing target RNAs and the m^6^A modification of OAS3 mRNA. These novel insights enhance our understanding of the molecular mechanisms underlying tumor immune evasion and suggest that targeting OAS3 could be a promising strategy for augmenting CD8^+^ T cell responses against tumors.

## Results

### PC cells promote the polarization of Mφ towards IL-10^high^VEGF-A^high^ M2d

The presence of CD163^+^ M2 cells in the PC microenvironment was identified through IF staining on tissue slices. The results revealed a significantly higher infiltration of CD163^+^ M2 cells in tumors compared to normal tissue. The mean fluorescence intensity (MFI) of CD163 was measured as 3.07 and 7.10 per high-power field (HPF) in normal and tumor tissues at stages II-III, respectively. Moreover, an upregulation in the level of CD163^+^ M2 cells was observed in stage Ⅳ, with MFIs of CD163 at 3.08 and 15.0 per HPF, respectively (**Fig. 1A**). To investigate whether PC cells promote M2 cell polarization, we first induced THP-1 cells into macrophages (Mφ) using PMA. Subsequently, we co-cultured these cells with Hpne and human PC cells (Panc-1, Bxpc-1, Aspc-1) using a transwell co-culture system for 48 hours. The results showed that co-culture with PC cells significantly promoted the polarization of CD11b^+^CD163^+^ M2 cells, with Panc-1 exhibiting the most pronounced effect (**Fig. 1B**). PCR analysis further demonstrated a significant upregulation of M2 markers ARG-1, CD163, and CD206 at the mRNA level after co-culture with PC cells, confirming the ability of PC cells to promote M2 polarization. Interestingly, among the M2-secreted cytokines, only IL-10 and VEGF-A showed significant increases, leading us to speculate that PC cells mainly drive Mφ polarization towards IL-10^high^VEGF-A^high^ M2d (**Fig. 1C-D**). Single-cell sequencing results revealed a higher infiltration of M2 cells, particularly M2d, in PC tumors compared to M1 (**Fig. 1E-F**). Additionally, an analysis of immune cell distribution in normal pancreas and PC tissues showed a significant increase in macrophage infiltration in tumor tissues (**Fig. S1A-B**). The proportion of Treg cells also increased, while the proportion of CD8^+^ T cells significantly decreased (**Fig. S1C-D**). Temporal analysis indicated that with disease progression, M2 cells predominantly polarized towards the M2d (**Fig. S1E-F**).

**Figure 1.**
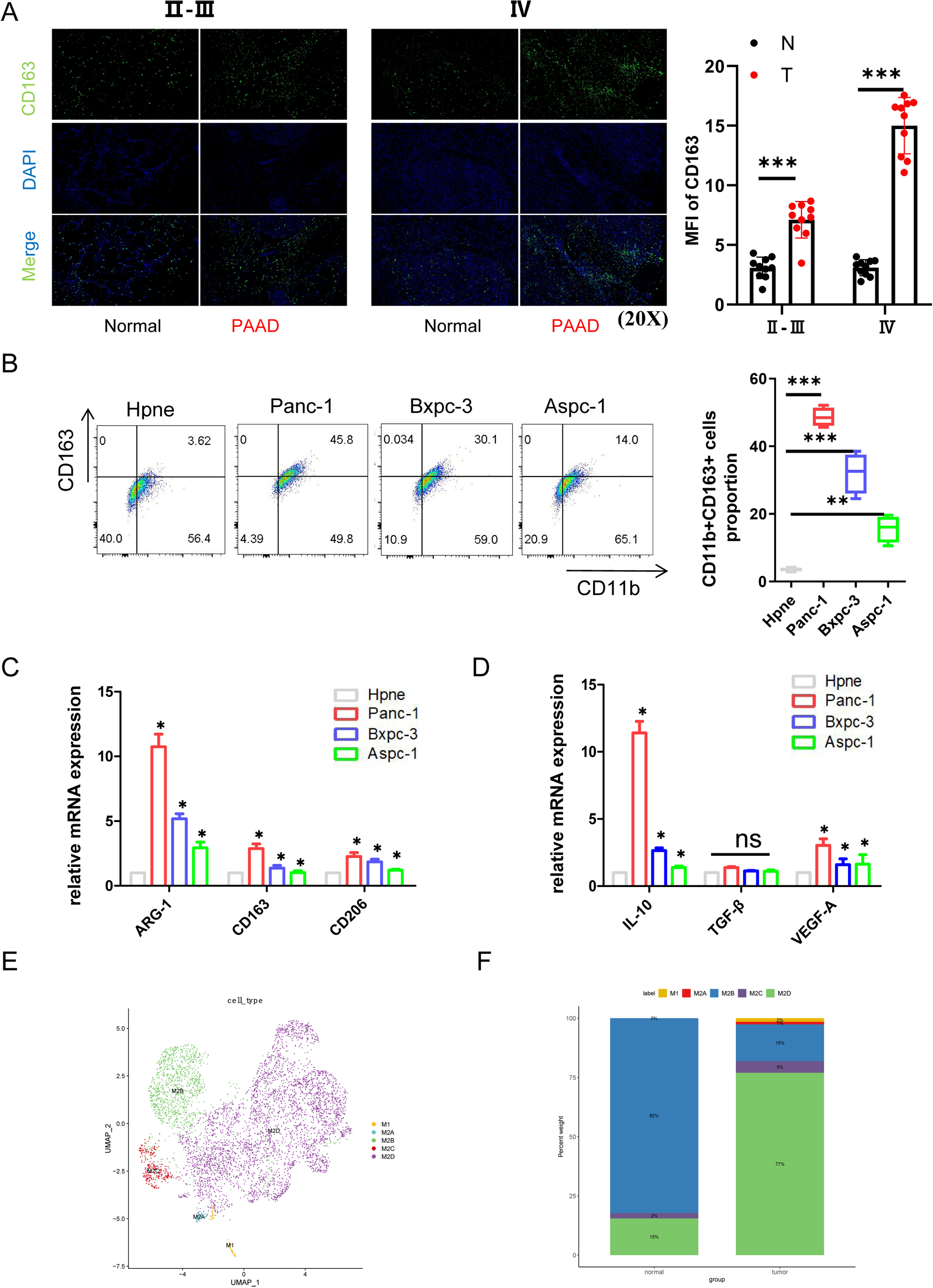
Pancreatic cancer cells promote the polarization of Mφ towards IL-10^high^VEGF-A^high^ M2d. A. Left, IF staining of human pancreatic cancer (PC) patient samples at different stages for CD163. Right, quantification of CD163^+^ macrophages per 20x microscopic field in tissue sections (n=10). B. Left, Mφ were co-cultured with Hpne cells and human pancreatic cancer cells (Panc-1, Bxpc-3, Aspc-1) using a transwell system for 48 hours, followed by flow cytometry analysis of the proportion of CD11b^+^CD163^+^ M2. Right, proportion of M2d in different groups (n=3). C. Relative mRNA expression of M2 markers ARG-1, CD163, and CD206 in Mφ after co-culture with Hpne and human pancreatic cancer cells. D. Relative mRNA expression of IL-10, TGF-β, and VEGF-A in Mφ after co-culture with Hpne and human pancreatic cancer cells. E. The UMAP plot of macrophages grouped into 5 phenotypes. F. Percentage of each distinct macrophages in the tumor and normal samples. Statistical significance was determined by Student t test. *, *P* < 0.05, **, *P* < 0.01, ns., no significance.

### PC cells secrete high levels of lactate, promoting the polarization of immunosuppressive M2d

To verify whether M2d induced by co-culture with PC cells possess immunosuppressive functions, we added lymphocytes extracted from human peripheral blood into M2d, after 48 hours of co-culture, we used flow cytometry to measure the secretion levels of GZMB and TNF-α in CD8^+^ cells. The results showed that the immunosuppressive capability of M2d in inhibiting CD8^+^ cell function was significantly higher than that of untreated macrophages (**Fig. 2A**). Hypoxia and high lactate levels are characteristics of the pancreatic cancer TME. We found that the lactate concentration in the supernatant of PC cell cultures was higher than that in Hpne cultures. Additionally, the intracellular lactate concentration in macrophages co-cultured with PC cells also significantly increased (**Fig. 2B**), suggesting that lactate may play a role in M2d polarization. To test this hypothesis, we used Panc-1 cells, which had the highest lactate concentration, for further study. We pretreated Panc-1 cells with 2-DG for 24 hours to inhibit lactate synthesis, or used the lactate transport inhibitor BAY-8002 to reduce lactate uptake in macrophages, thus lowering intracellular lactate levels (**Fig. 2C**). PCR analysis revealed that, compared to TCM, the mRNA levels of M2d markers ARG-1, CD163, CD206, IL-10, and VEGF-A were significantly reduced in the two treated groups (**Fig. 2D**). IF staining indicated that 2-DG and BAY-8002 treatments inhibited the promotion of M2d polarization by Panc-1 co-culture (**Fig. 2E**). Flow cytometry analysis also demonstrated that Panc-1 co-culture promoted CD11b^+^CD163^+^ M2d polarization, whereas inhibition of lactate secretion or transport by 2-DG or BAY-8002 decreased the proportion of M2d (**Fig. 2F**). To more directly investigate the role of lactate in M2d polarization, we added lactate to macrophage cultures in normal 1640 medium or in TCM and setting up a concentration gradient. The results confirmed that the addition of lactate promoted M2d polarization in both normal medium and TCM groups, though the effect quickly saturated in the TCM group (**Fig. S2A**). Interestingly, we found that the addition of TCM from Panc-1 cancer cells inhibited apoptosis in macrophages under hypoxic conditions, whereas the apoptosis level of CD86^+^ M1 macrophages significantly increased (**Fig. S2B**). This might be another potential mechanism through which PC cells promote M2d polarization.

**Figure 2.**
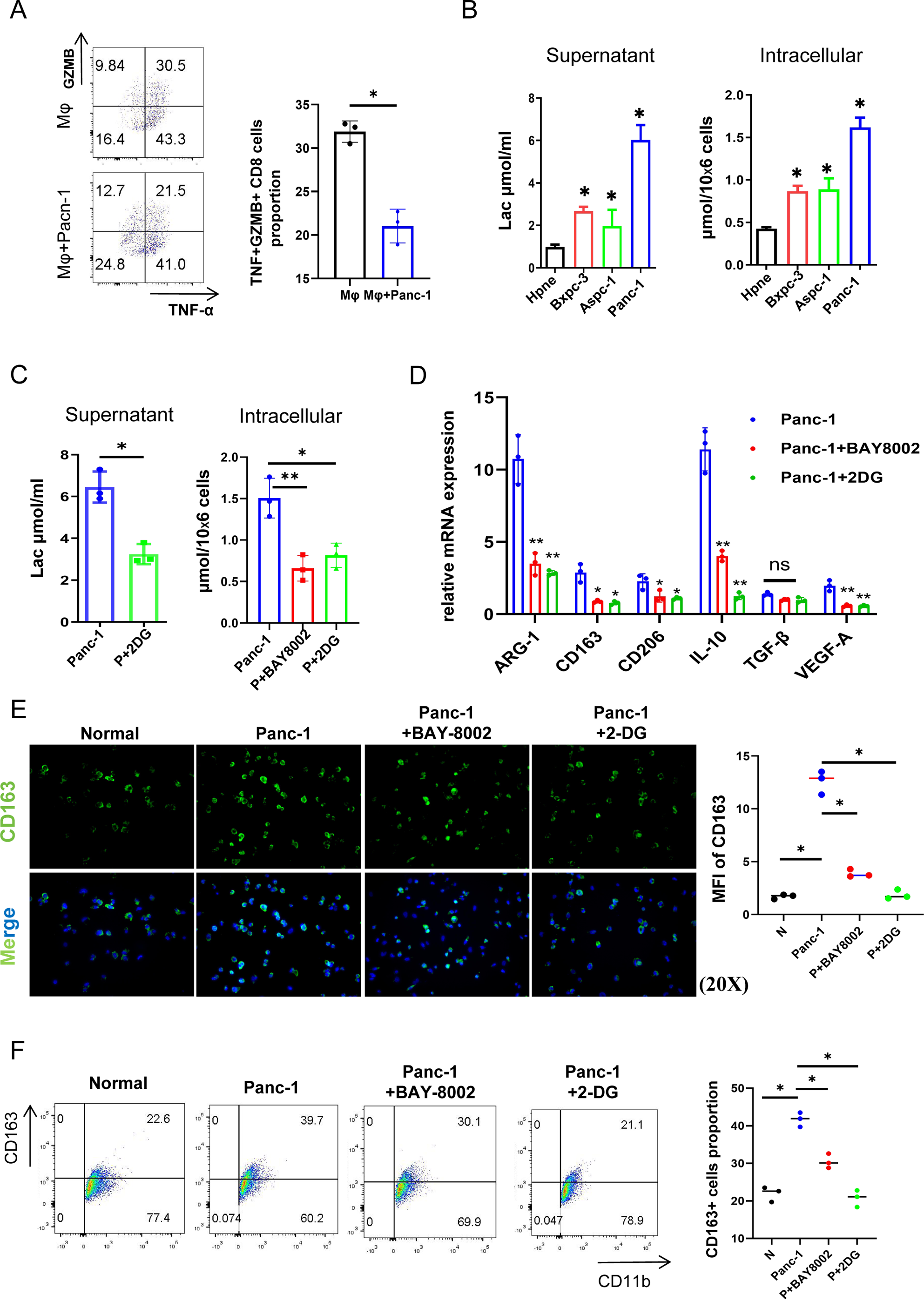
Pancreatic cancer cells secrete high level of lactate, promoting the polarization of M2d. A. Left, flow cytometry analysis of the inhibition of Mφ on granzyme B (GZMB) and TNF-α secretion of CD8^+^ T cell. Right, proportion of TNF^+^GZMB^+^CD8^+^ T cells in different groups (n=3). B. Left, lactate concentration in the culture supernatant of Hpne and pancreatic cancer cells. Right, intracellular lactate concentration in Mφ after co-culture with different cells (n=3). C. Left, The lactate concentration in the tumor-conditioned medium (TCM) from Panc-1 cells pretreated with 2-DG for 24 hours. Right, The intracellular lactate concentration in Mφ treated with Panc-1 TCM, Panc-1 TCM+BAY8002, and Panc-1+2DG TCM (n=3). D. Relative mRNA expression of M2 markers ARG-1, CD163, and CD206, along with immune factors IL-10, TGF-β, and VEGF-A, in different groups (n=3). E. Left: IF analysis showing the levels of CD163 in macrophages across different treatment groups. Right: Mean fluorescence intensity (MFI) of CD163 in Mφ in different groups (n=3). F. Left: Flow cytometry analysis showing the polarization levels of CD11b^+^CD163^+^ M2 cells in different treatment groups. Right, proportion of CD11b^+^CD163^+^ M2 in different groups (n=3). Statistical significance was determined by Student t test. *, *P* < 0.05, ns., no significance.

### Lactate promotes M2d polarization by upregulating METTL3 in macrophages

To further investigate the mechanism by which lactate promotes M2d polarization, we performed RNA-seq analysis on macrophages co-cultured with Panc-1 cells, 2-DG pretreated Panc-1 cells or normally cultured. Differential gene expression analysis using disease ontology (DO) revealed a significant enrichment of pancreatic cancer-related signaling pathways in macrophages after co-cultured with Panc-1 cells (**Fig. 3A**). Following the inhibition of lactate secretion by 2-DG, gene ontology (GO) enrichment analysis suggested that lactate-promoted M2d polarization is closely associated with RNA biosynthesis and metabolic processes (**Fig. 3B**). RNA N6-methyladenosine (m^6^A) modification, the most abundant modification in mRNA, is recognized as a key layer of the epiregulome. METTL3, a critical enzyme for m6A modification, was found to be downregulated following the inhibition of lactate synthesis in Panc-1 cells (**Fig. 3C**), while other modifying enzymes showed no significant differences (data not shown). Western blot analysis revealed a significant increase in METTL3 protein expression in macrophages after co-culture with PC cells (**Fig. 3D**). To further clarify the regulatory effect of lactate on METTL3, we examined METTL3 protein expression levels in macrophages under different lactate concentrations, confirming that lactate upregulates METTL3 (**Fig. 3E**). IF staining of clinical pathological tissues also showed that infiltrating M2 cells expressed high levels of METTL3 (**Fig. 3F**). Areas with high M2 cell infiltration also exhibited increased METTL3 expression, with a clear co-localization phenomenon (**Fig. S3A**). Therefore, we hypothesize that PC cells may promote M2d polarization by secreting lactate to upregulate METTL3 expression in macrophages. To test this hypothesis, we constructed METTL3-knockdown THP-1 cell lines (**Fig. S3B**). Flow cytometry analysis showed that the ability of Panc-1 TCM to promote M2 polarization was significantly reduced following METTL3 knockdown, with no significant difference from the control group (**Fig. 3G**). IF staining further confirmed that the proportion of M2d polarization decreased significantly in METTL3-knockdown macrophages (**Fig. 3H**).

**Figure 3.**
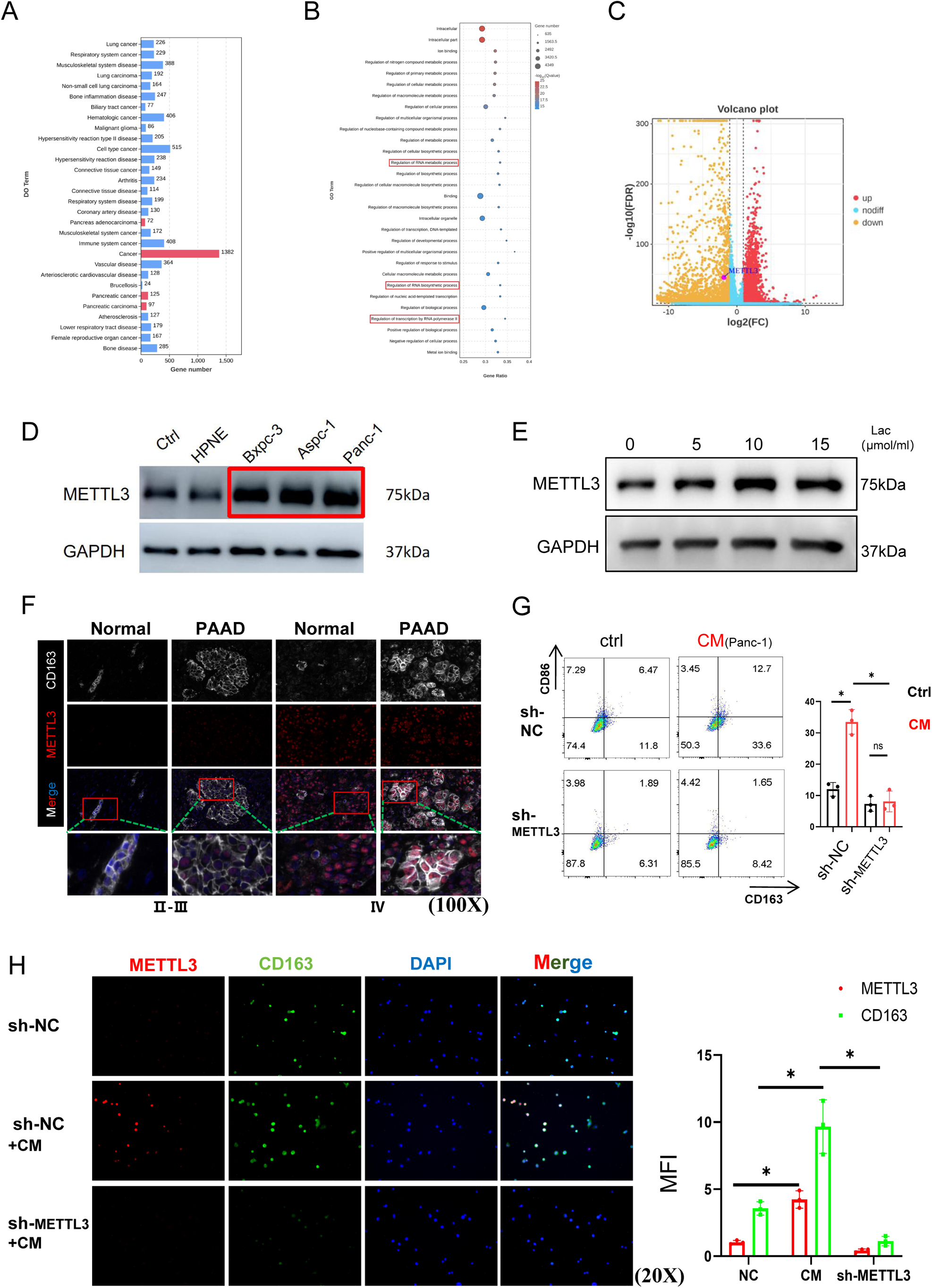
Lactate promotes M2d polarization by upregulating METTL3 in macrophages. A and B. Disease ontology (DO) terms (A) and gene ontology (GO) terms (B) of the differentially expressed genes in TCM-treated macrophages compared with TCM+2DG-treated macrophages. C. Volcano plot of differentially expressed genes indicates higher expression of METTL3 in TCM-treated macrophages compared to TCM+2DG-treated macrophages. D. WB images showing the expression of METTL3 in macrophages after co-culture with normal 1640 medium, Hpne cells, and human pancreatic cancer cells (Panc-1, Bxpc-3, Aspc-1) for 48 hours. E. Western blot images showing the expression of METTL3 in macrophages under different lactate concentration treatments. F. IF staining showing the colocalization of CD163 and METTL3 in human PC patient samples at different stages. G. Left: Flow cytometry analysis showing the polarization level of CD163^+^CD86^−^ M2 cells in sh-METTL3 macrophages under normal medium and CM culture conditions. Right: Proportion of CD163^+^CD86^−^ M2 cells in different groups (n=3). H. Left: IF analysis showing the expression levels of CD163 and METTL3 in macrophages across different treatment groups. Right: MFI of CD163 and METTL3 in macrophages in different groups (n=3). Statistical significance was determined by Student t test. *, *P* < 0.05, **, *P* < 0.01, ns., no significance.

### METTL3 plays a pivotal role in preserving the immunosuppressive phenotype of macrophages

PD-L1 is a crucial immune inhibitory protein that plays a significant role in the immunosuppressive function of M2. Flow cytometry analysis revealed that PD-L1 expression levels in CD11b^+^CD163^+^ M2 decreased following METTL3 knockdown, suggesting that METTL3 not only regulates M2 polarization but also plays an important role in their immunosuppressive function (**Fig. 4A**). Furthermore, ELISA results showed that the secretion levels of IL-10 and VEGF-A in the supernatant of macrophages were significantly increased after 48 hours of stimulation with Panc-1 TCM. However, when using sh-METTL3 macrophages or reducing intracellular lactate levels by 2-DG or BAY-8002, the levels of IL-10 and VEGF-A exhibited a decreasing trend (**Fig. 4B-C**). IL-6, an important inflammatory cytokine recently implicated in promoting PC development and progression, was found to be highly secreted by macrophages stimulated with Panc-1 TCM. This secretion was inhibited by METTL3 knockdown and lactate reduction, consistent with the previous findings (**Fig. 4D**). These results may partially explain why PC, characterized by hypoxia and high lactate levels, often exhibits poor response to immunotherapy. Other key immune factors were also examined; IL-1β, another critical pro-inflammatory cytokine, showed similar trends as IL-6 (**Fig. S4A**), while anti-inflammatory factor TGF-β and tumor-killing factor IFN-γ showed no significant differences among the groups (**Fig. S4B-C**). Granzyme B (GZMB) and tumor necrosis factor-alpha (TNF-α) are essential mediators of the cytotoxic activity of CD8^+^ T cells. Our study found that the ability of macrophages stimulated with Panc-1 TCM to suppress GZMB and TNF-α secretion by CD8^+^ T cells was significantly weakened following METTL3 knockdown (**Fig. 4E**). VEGF-A is a key factor promoting tumor angiogenesis, we extracted the supernatants from macrophages and sh-METTL3 macrophages stimulated with TCM and added them to human umbilical vein endothelial cells (HUVECs) for a tube formation assay. The results showed that the tube formation area was significantly reduced in the sh-METTL3 group (**Fig. 4F**). These experiments further clarified the critical role of METTL3 in PC cell-induced IL-10^high^VEGF-A^high^ M2d polarization, CD8^+^T cell immune suppression, and promotion of angiogenesis.

**Figure 4.**
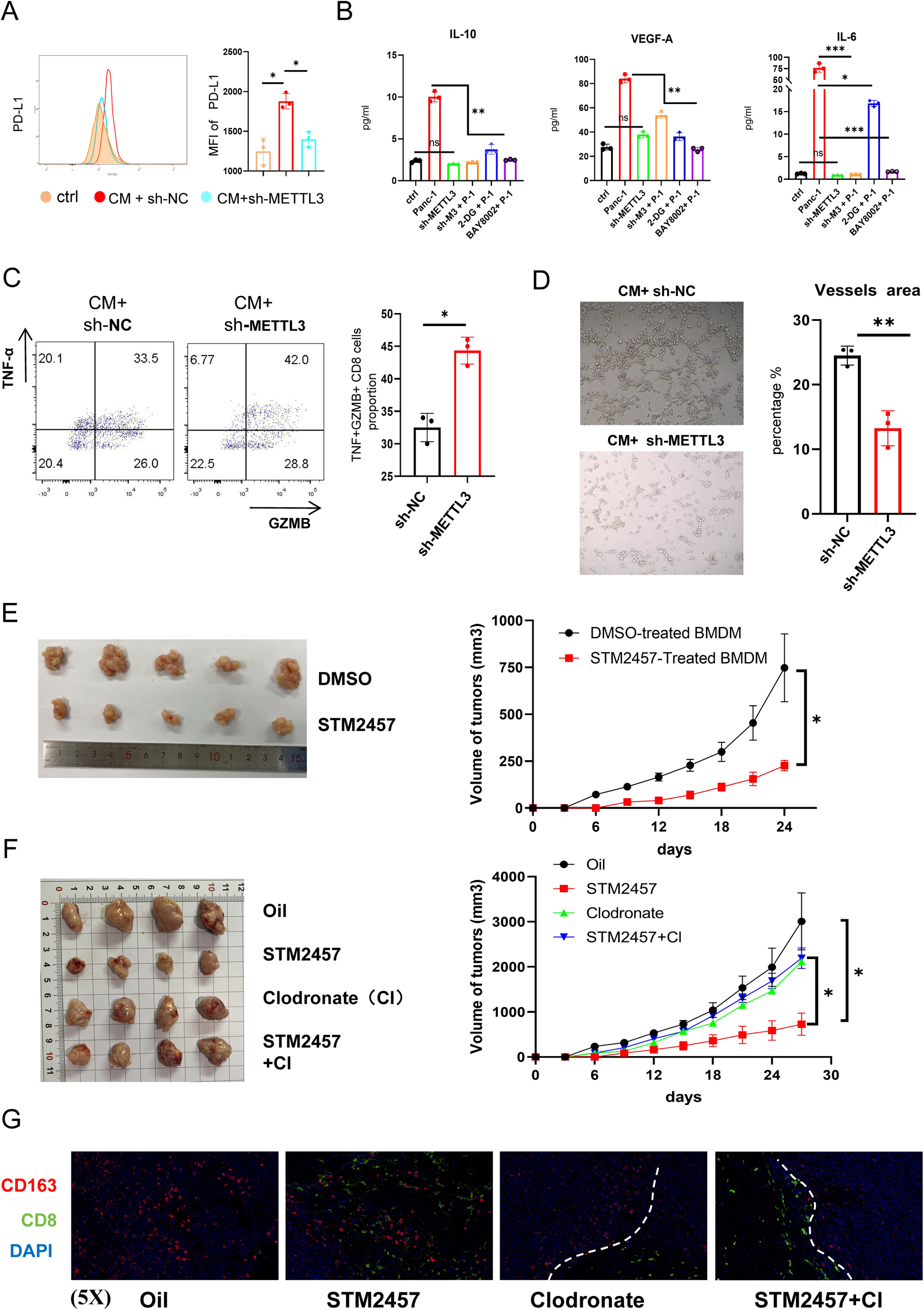
METTL3 plays a pivotal role in preserving the immunosuppressive function of TAMs. A. Left: Flow cytometry analysis showing the effect of CM on PD-L1 in sh-NC Mφ and sh-METTL3 Mφ. Right: Proportion of PD-L1^+^ Mφ (n=3). B-D. ELISA measurements of IL-10 (B), VEGF-A (C), and IL-6 (D) release under different culture conditions in sh-NC Mφ and sh-METTL3 Mφ (n=3). E. Left: Flow cytometry analysis showing the effect of METTL3 knockdown in Mφ on the secretion of GZMB and TNF-α by CD8^+^ T cells under CM culture conditions. Right: Proportion of TNF^+^GZMB^+^CD8^+^ T cells in different groups (n=3). F. The effect of TAMs culture supernatants from different treatment groups on the angiogenic capacity of HUVECs (n=3). G-H. DMSO or STM2457 pre-treated BMDMs mixed with PANC-02 cells were inoculated subcutaneously into Balb/c-Nude mice. Images of excised tumors and tumor growth curves were shown (n=5). H-J. PANC-02 cells were inoculated subcutaneously with oil, STM2457, or Clodronate into pre-treated C57BL/6J mice (n=4). K. IF analysis showing CD163^+^ M2 and CD8^+^ T cell infiltration in tumor tissues from C57BL/6J mice. Statistical significance was determined by Student t test. *, *P* < 0.05; **, *P* < 0.01, ***, *P* < 0.001, ns., no significance.

To further validate the immunoregulatory role of METTL3 expression in macrophages *in vivo*, we designed two pancreatic cancer mouse model experiments. First, we extracted bone marrow-derived macrophages (BMDMs) from mice and pre-treated them with either DMSO or STM2457 (a METTL3 inhibitor, 30 mg/kg). These pre-treated BMDMs were then mixed with PANC-02 cells and injected subcutaneously into Balb/c-Nude mice. Monitoring the subcutaneous tumor volumes revealed that inhibiting METTL3 expression in BMDMs significantly slowed tumor growth (**Fig. 4G-H**). To demonstrate that whether PC cells can promote the formation of the tumor immune microenvironment by upregulating METTL3 in macrophages, we pre-treated C57 mice with intraperitoneal injections of corn oil, STM2457, and Clodronate (macrophage depletion agent). Subsequently, we established subcutaneous PANC-02 tumor models in these mice. Comparisons showed that intraperitoneal injection of STM2457 inhibited tumor growth, but this inhibitory effect was markedly reduced when macrophages were also depleted (**Fig. 4I-J**). Interestingly, IF analysis of tumor tissues revealed decreased CD163^+^M2 and increased CD8^+^ T cells in the STM2457 group. Although some CD8^+^ T cell infiltration was observed in the Clodronate and STM2457 + Clodronate groups, these cells appeared to be confined to the tumor stroma and did not infiltrate the tumor tissue itself (**Fig. 4K**). These results highlight the crucial role of METTL3 in macrophages in promoting PC progression and shaping the tumor immune microenvironment.

### METTL3 promotes M2d polarization by upregulating OAS3 through methylation modification

METTL3 is a key protein mediating m^6^A modifications involved in the RNA methylation process. Using RNA-seq, we identified 62 genes with significantly upregulated mRNA levels after CM treatment but reduced or even eliminated upon METTL3 knockdown or decreased CM lactate concentration (**Fig. 5A**). Analysis of the top ten upregulated genes in Mφ after co-cultured with PC cells revealed that 2’-5’-oligoadenylate synthase 3 (OAS3) were negatively correlated with PC prognosis (**Fig. 5B**, **Fig. S5A**). We used MeRIP-seq (Methylated RNA Immunoprecipitation Sequencing) to assess the changes in methylation levels of the top ten genes in and found that OAS3 had two sites with significantly decreased methylation upon METTL3 knockdown (**Fig. 5C**). As previously noted, the mRNA level changes of OAS3 were highly correlated with METTL3 and lactate levels (**Fig. 5D**). WB analysis revealed that Panc-1 CM treatment upregulated OAS3 protein levels, METTL3 knockdown or decreased CM lactate concentration resulted in significantly reduced OAS3 protein levels (**Fig. 5E**). Additionally, OAS3 protein expression exhibited a clear lactate concentration dependency (**Fig. 5F**), suggesting the regulation of OAS3 expression via lactate/METTL3 axis. The TCGA dataset revealed that OAS3 expression is significantly elevated in PC tissues compared to normal tissues (**Fig. S5B**). Additionally, we observed higher immune and stromal scores in the OAS3 high-expression group (**Fig.S5C-D**). Previous studies indicated that immune-excluded tumors are characterized by abundant immune cells that do not penetrate the tumor parenchyma but rather accumulate around the peritumoral stroma(21). Consequently, we hypothesized that the rich stromal components (such as TAMs) in the pancreatic tumors of the OAS3 high-expression group may inhibit the potential anti-tumor immune response. Supporting our hypothesis, our ssGSEA results identified significantly higher levels of TAM infiltration in the OAS3 high-expression group (**Fig. S5E**). Furthermore, our analysis also showed a significant positive correlation between OAS3 expression and markers of T-cell exhaustion and immune checkpoints, suggesting that OAS3 may be involved in T-cell exhaustion leading to immune escape within the PC TME (**Fig. S6A-B**). Therefore, we proposed that OAS3 might serve as a critical target through which METTL3 regulates TAM infiltration, T-cell exhaustion, and promotes tumor progression.

**Figure 5.**
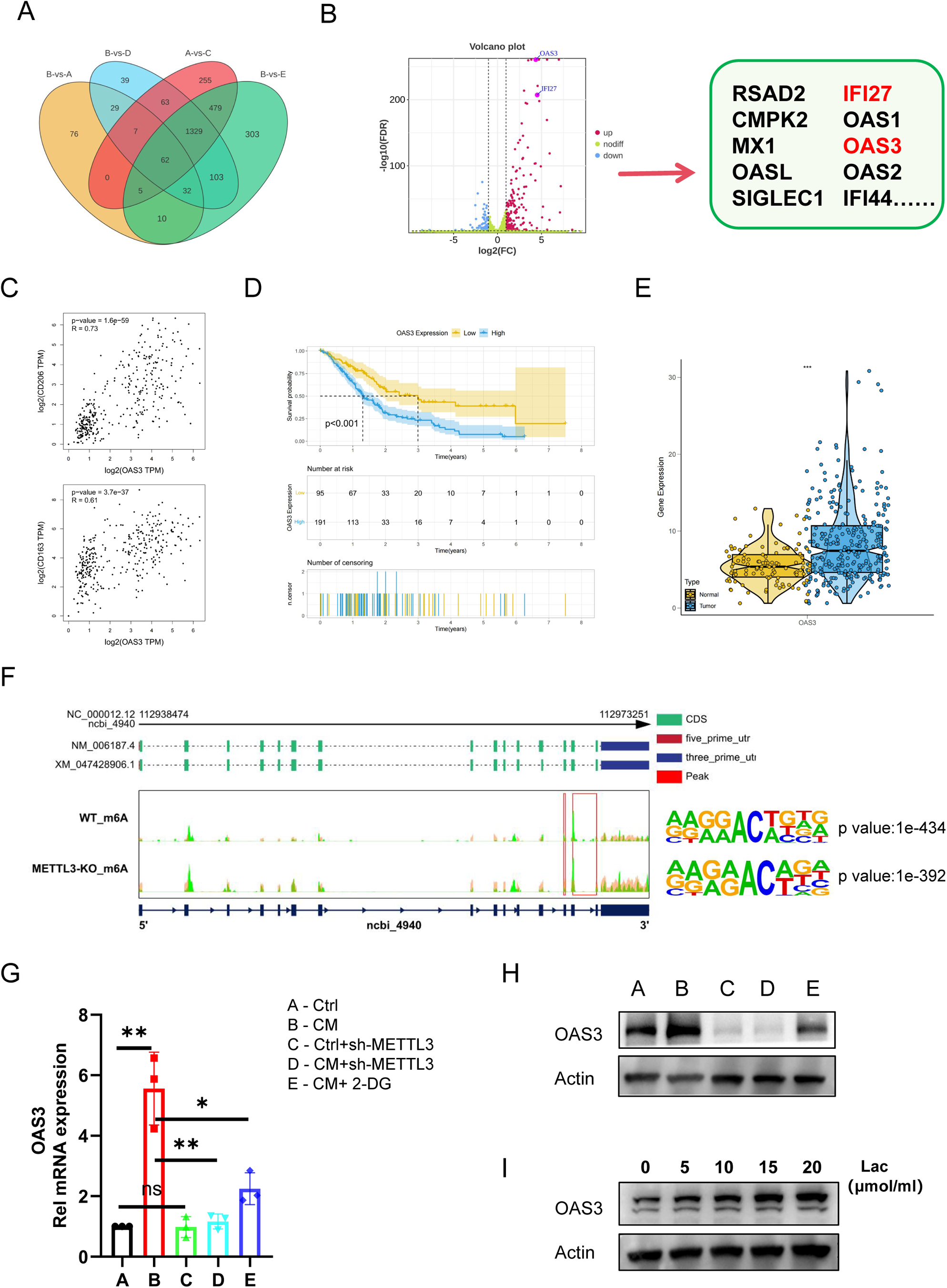
METTL3 promotes M2d polarization by upregulating OAS3 through methylation modification. A.Venn diagram showing the differential gene expression between different cell groups detected by RNA-seq. B. Volcano plot displaying the top ten upregulated genes in Mφ after co-culture with cancer cells (n=3). C. Scatter plots showing positive correlations between OAS3 expression and CD163\CD206. D.Kaplan–Meier curves for the OAS3 high and low expression groups based on 286 PDAC patients from the TCGA-cohort, including 95 samples in low group and 191 samples in high group (log-rank test). The low group showed a significantly better prognosis. E. The difference of mRNA expression level of OAS3 between normal and tumor TCGA-PDAC samples. F. Integrative genomics viewer (IGV) tracks showing m^6^A peaks distributed on the OAS3 transcript from MeRIP-seq data in Mφ. G. qRT-PCR analysis of the relative mRNA levels of OAS3 in different treatment groups of macrophages and sh-METTL3 macrophages (n=3). H. Western blot images showing the expression of OAS3 in different treatment groups of macrophages and sh-METTL3 macrophages. I. Western blot images showing the expression of OAS3 in macrophages under different lactate concentration treatments. Statistical significance was determined by Student t test. ***, *P* < 0.001.

To confirm this hypothesis, we constructed sh-NC and sh-OAS3 THP-1 cell lines and validated the knockdown efficiency (**Fig. 5G**). Subsequently, we induced sh-NC Mφ and sh-OAS3 Mφ cells and stimulated their polarization using either normal culture medium or Panc-1 conditioned medium (CM). Flow cytometry analysis revealed that OAS3 knockdown significantly inhibited the CM-induced polarization of Mφ cells towards the CD163^+^CD86^−^ M2 (**Fig. 5H**). This phenomenon was further confirmed by IF detection of CD163 expression levels (**Fig. 5I**). Through these experiments, we preliminarily verified that METTL3 promotes M2d polarization via methylation-mediated upregulation of OAS3. This mechanism is a potential pathway by which PC cells create an immunosuppressive TME, thereby facilitating tumor progression.

### Specific OAS3 inhibition in macrophage enhances the efficacy of gemcitabine and anti-PD-L1 mAb treatment in PC

To further investigate whether OAS3 influences PD-L1 expression in M2d, we divided macrophages into three groups: A: ctrl+sh-NC Mφ, B: CM+sh-NC Mφ and C: CM+sh-OAS3 Mφ. After 48 hours of stimulation, flow cytometry analysis revealed that OAS3 knockdown significantly reversed the CM-induced upregulation of PD-L1 in macrophages (**Fig. 6A**). Concurrently, IL-10 and VEGF-A secretion levels were also significantly reduced in the sh-OAS3 Mφ group (**Fig. 6B-C**). Based on these findings, we hypothesized that reduced IL-10 secretion following OAS3 knockdown could alleviate the suppressive effect of M2 on CD8^+^T cells. To validate this hypothesis, we maintained the three-group division and stimulated the cells for 48 hours. After replacing the supernatant, lymphocytes isolated from the peripheral blood of healthy volunteers were added to the cell culture system for co-culture for another 48 hours. Flow cytometry analysis revealed a significant increase in the proportion of CD8^+^T cells in the CM+sh-OAS3 Mφ group. More importantly, the proportion of TNF^+^GZMB^+^ CD8^+^ T cells also significantly increased (**Fig. 6D**). Similarly, tube formation assays demonstrated that the angiogenic function of M2 was markedly inhibited following OAS3 knockdown (**Fig. 6E**). Through these experiments, we have further elucidated that OAS3 knockdown not only reduces PD-L1 expression but also diminishes the immunosuppressive and pro-angiogenic capabilities of M2d, thereby restoring CD8^+^T cell activity and potentially improving antitumor immune responses.

**Figure 6.**
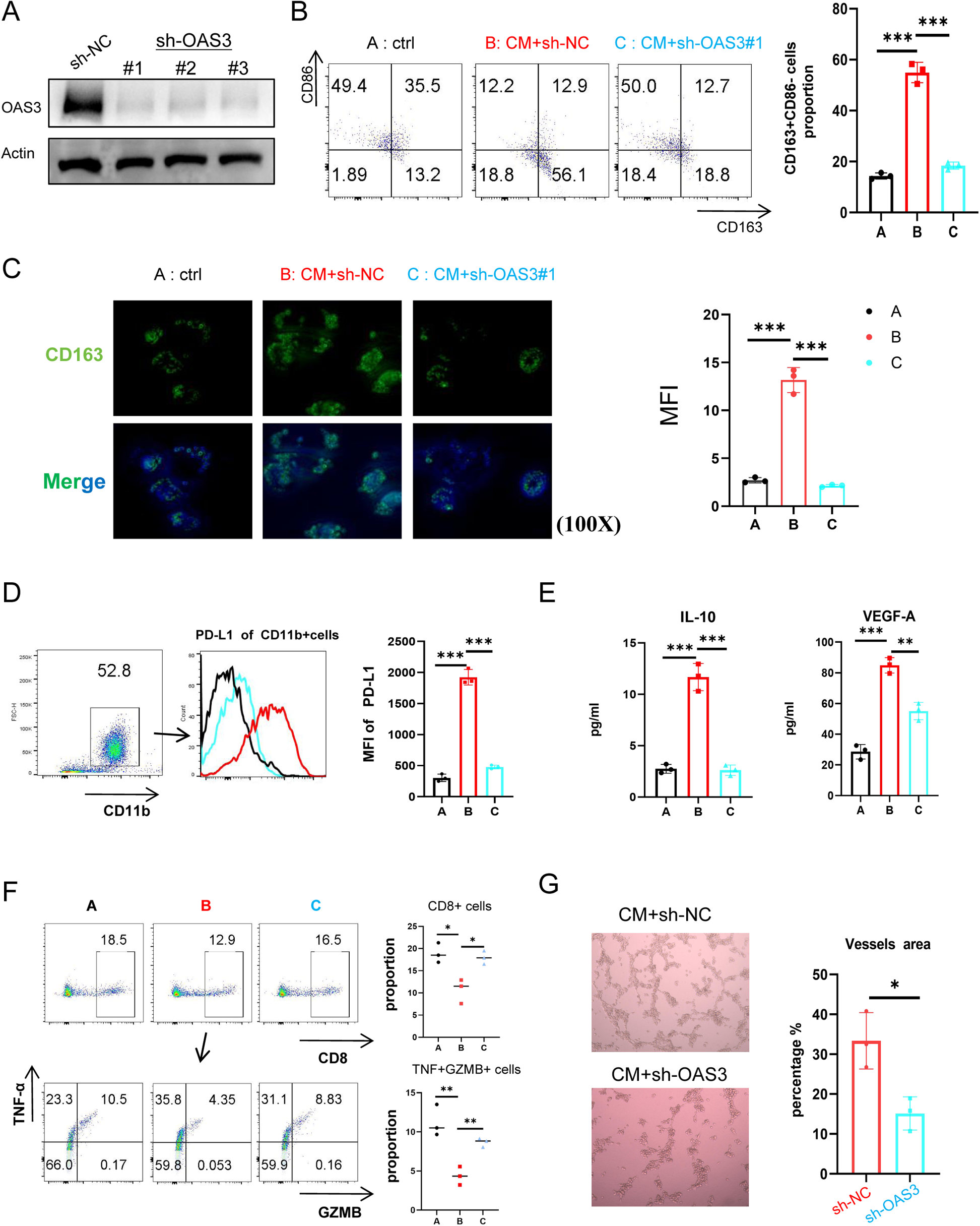
Specific OAS3 inhibition in macrophage impaird M2 immunosuppressive function *in vitvo*. A. Western blot images verifying the knockdown effect of OAS3. B. Left: Flow cytometry analysis showing the polarization level of CD163^+^CD86^−^ M2 cells in sh-NC and sh-OAS3 Mφ under normal or CM conditions. Right: Proportion of CD163^+^CD86^−^ M2 cells in different groups (n=3). C. IF analysis showing the expression levels of CD163 in Mφ treated with/without CM (n=3). D. Flow cytometry analysis showing PD-L1 expression levels in sh-NC or sh-OAS3 Mφ under different culture conditions. E. ELISA measurements of IL-10 (left) and VEGF-A (right) secretion levels in sh-NC and sh-OAS3 Mφ under different culture conditions (n=3). F. Flow cytometry analysis showing the effect of OAS3 knockdown in Mφ on the secretion levels of GZMB and TNF-α by CD8^+^ T cells under different culture conditions. Right: Proportion of TNF^+^GZMB^+^CD8^+^ T cells in different groups (n=3). G. The effect of culture supernatants from Mφ or sh-OAS3 Mφ on the angiogenic capacity of HUVECs (n=3). Statistical significance was determined by Student t test. *, *P* < 0.05; **, *P* < 0.01, ***, *P* < 0.001.

To investigate the impact of OAS3 protein in macrophages on immune regulation and tumor immunotherapy efficacy *in vivo*, we constructed a humanized mouse model by intraperitoneal injection of human PBMCs. 4 and 8 weeks post-injection, human CD3^+^CD4^+^ T cells and CD3^+^CD8^+^ T cells were successfully detected in the peripheral blood of the mice, confirming the model’s validity. We utilized Panc-1 cells to construct a pancreatic cancer cell line-derived xenograft (CDX) model at 5^th^ week (**Fig. S7A-B**). The mice were then divided into four groups. Groups I and III received peritumoral injections of sh-NC Mφ, while groups II and IV received sh-OAS3 Mφ (every 3 days). When palpable tumors were observed subcutaneously, groups I and II received intraperitoneal injections of corn oil (100 µL/mouse), whereas groups III and IV received intraperitoneal injections of Gem (20 mg/kg) and PD-L1 mAb (10 mg/kg) three times per week. At 9^th^ week, the mice were euthanized, and tumors were excised following the collection of blood from the retro-orbital sinus. Images revealed that tumors in the sh-OAS3 Mφ peritumoral injection group were significantly smaller than those in the sh-NC Mφ group. Notably, specific knockdown of OAS3 in Mφ, combined with Gemand PD-L1 mAb treatment, demonstrated a synergistic effect (**Fig. 6F**). Flow cytometry analysis of CD8^+^ T cells in peripheral blood indicated that peritumoral injection of sh-OAS3 Mφ increased the proportion of CD8^+^ T cells, particularly when combined with immunotherapy, showing an even more pronounced effect (**Fig. 6G**). IF staining showed that the stromal regions of tumors in the sh-OAS3 group exhibited significantly thinner and incomplete encapsulation by CD163^+^M2, along with a modest infiltration of CD8^+^T cells. Treatment with Gem and PD-L1 mAb did not improve infiltrated CD8^+^T cells obviously, but OAS3 knockdown resulted in a synergistic effect, significantly increasing the infiltration of CD8^+^ T cells within the tumors (**Fig. 6H**). Through these experiments, we demonstrated that OAS3 knockdown in macrophages enhances the efficacy of tumor immunotherapy by increasing CD8^+^ T cell infiltration and reducing tumor growth, particularly when combined with Gem and PD-L1 blockade. These results partially demonstrate that targeting OAS3 inhibitors have potential clinical value in improving PC immunotherapy.

**Figure 7.**
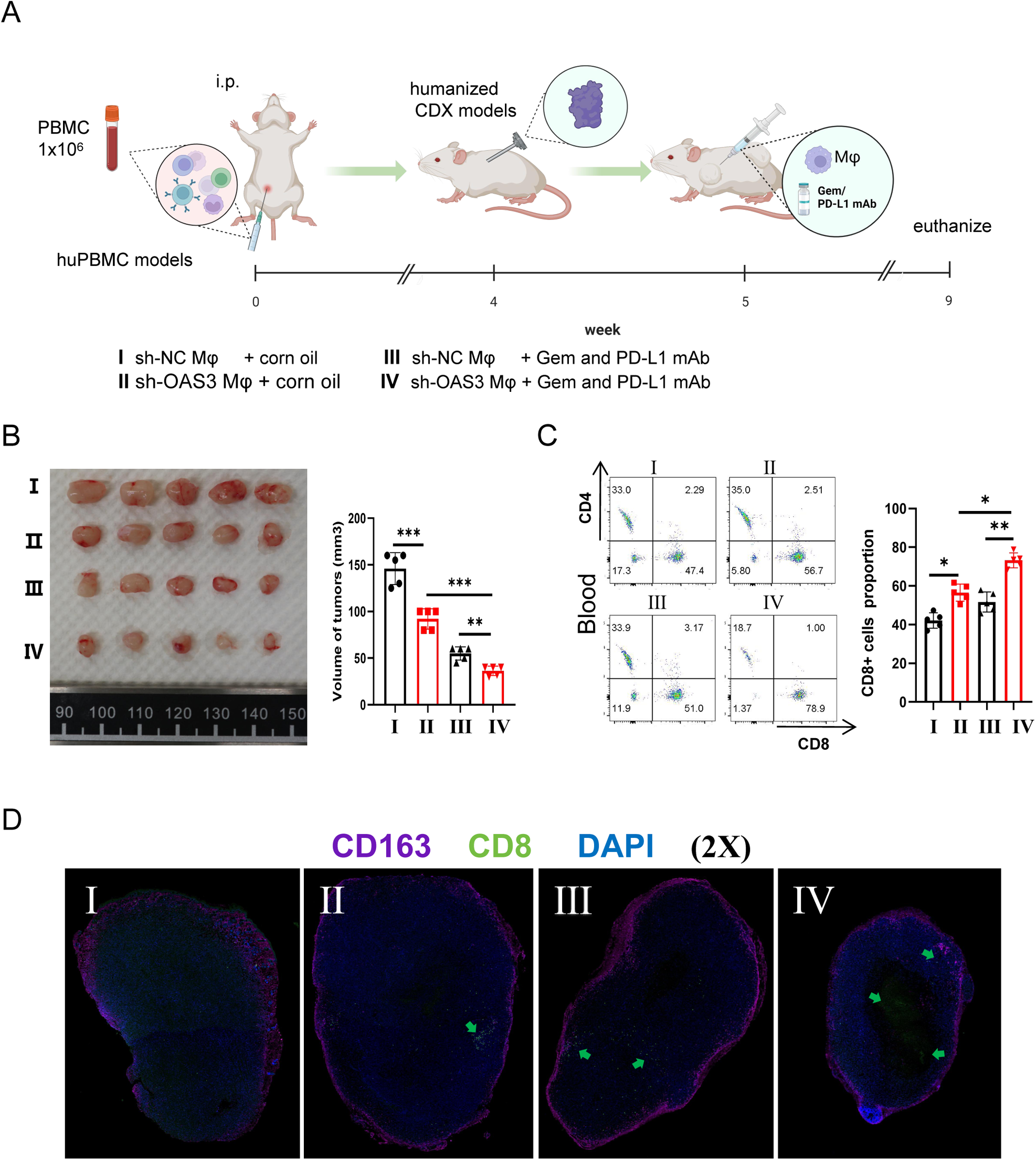
Specific OAS3 inhibition in macrophage enhances the efficacy of gemcitabine and anti-PD-L1 mAb treatment in PC *in vivo*. A. Intraperitoneal injection of human PBMCs to establish a humanized mouse model of the immune system and construction of a pancreatic cancer CDX model. B. In the pancreatic cancer CDX (Panc-1) model, intraperitoneal injection of gemcitabine and PD-L1 mAb or peritumoral injection of macrophages; excised tumor images and tumor growth curves are shown (n=5). C. Flow cytometry analysis showing the changes in the proportion of CD8^+^ T cells in peripheral blood of CDX (Panc-1) model mice from different groups (n=5). D. IF analysis showing CD163^+^ M2 and CD8^+^ T cells infiltration in tumor tissues of CDX (Panc-1) model mice. Statistical significance was determined by Student t test. *, *P* < 0.05; **, *P* < 0.01, ***, *P* < 0.001.

## Discussion

In recent years, personalized strategies to improve PC treatment have been increasingly integrated into clinical practice, with particular attention to targeting TAMs. Our findings demonstrate that M2 infiltration in PC tissue is significantly higher than normal and increases with disease progression. Several approaches have been explored to target TAMs. The colony-stimulating factor 1 receptor (CSF1R) is crucial for macrophage survival, and monoclonal antibodies targeting CSF1R have shown an objective response rate of up to 71% in the treatment of tenosynovial giant cell tumors(22). The CCL2/CCR2 signaling axis mediates TAMs recruitment within the TME, and research indicates that targeting CCR2 can reduce TAMs infiltration, increase tumor-infiltrating lymphocytes and enhance tumor cytotoxicity(3). Additionally, phosphoinositide 3-kinase (PI3K) is highly expressed in myeloid cells, playing a vital role in tumor metastasis and immune evasion, while the overactivation of the signal transducer and activator of transcription type 3 (STAT3) pathway promotes M2 polarization(23). Tumor cells can express CD47, which binds to the SIRPα receptor on macrophages, transmitting a “don’t eat me” signal to inhibit antitumor activity and facilitate immune escape(24). Despite these advancements, TAM-targeting strategies have shown limited efficacy in PC treatment, highlighting the need for further exploration of TAMs regulatory mechanisms and novel targets.

TAMs mediate immune escape through various mechanisms, including secreting cytokines, expressing PD-L1, and recruiting immunosuppressive cells to the tumor site(25). In PC, TAMs can promote tumor progression in several ways: 1. inhibiting T cell proliferation, promoting their conversion to Th2 cells, and limiting their cytotoxicity(26). 2. Stimulated by PSCs, TAMs produce cytokines that activate myofibroblasts expressing α-SMA, contributing to fibrosis in the TME(9). 3. Secreting VEGF-A to promote tumor angiogenesis(27). 4. TAMs-derived IL-10 inhibits dendritic cell maturation, increases macrophage differentiation, reduces antigen presentation, inhibits IFN-γ release, prevents naive T cell differentiation and promotes immune escape(8). 5. TAMs-secreted TGF-β upregulates phosphorylation of PKM2 in PC cells and co-activates STAT1, ultimately increasing PD-L1 expression(10). In summary, TAMs infiltration in PC intensifies with disease progression, and their high-density infiltration is negatively correlated with overall patient survival.

Hypoxia and high lactate levels are significant features of the PC TME. We previously demonstrated that under hypoxia, PC cells can promote M2 polarization and subsequent metastasis via exosome-mediated delivery of miR-301a, which activates the PTEN/PI3Kγ/AKT/mTOR pathway(28). Lactate is another crucial mediator in PC cell-induced M2 macrophage polarization. G protein-coupled receptor 132 (Gpr132), a lactate sensor, is highly expressed in macrophages within the acidic TME. Loss of Gpr132 can inhibit M2 polarization and tumor metastasis(29). In PC, lactate is transported into macrophages via monocarboxylate transporters (MCT), stabilizing HIF-1α through NF-κB/p65 pathway and promoting M2 polarization. Targeting lactate dehydrogenase A (LDHA), inhibiting lactate transporters MCT1/4, or using 2-deoxyglucose (2DG) to inhibit glycolysis can suppress M2 polarization and enhance PC treatment efficacy(30). Hypoxia and high lactate levels have also been reported to synergistically activate the KRAS/MAPK pathway, promoting Arg1^+^ M2 cell polarization in PC. TAMs in hypoxic, high-lactate regions secrete higher levels of VEGF, promoting angiogenesis and tumor growth(13).

Advances in technology have highlighted the complexity and heterogeneity of macrophages, leading to the identification of new subpopulations and their respective functions. ARAS et al. classified TAMs into 7 categories based on specific markers, such as angiogenic (VEGF, ADM, PDGF), immunosuppressive (PD-L1, IL-10, TGF-β), mesenchymal-epithelial transition (TLR4/IL-10, TGF-β), and cancer stem cell interactions (TGF-β, IL-10, MFG-E8, IL-6)(31). TAMs are predominantly M2, which can be further divided into M2a, M2b, M2c, and M2d. M2c and M2d both inhibit immune responses and promote tumor growth. M2c, formed by stimulation with IL-10, glucocorticoids, and TGF-β, secrete TGF-β and IL-10 to promote tissue repair and matrix remodeling(12). M2d, stimulated by adenosine, IL-6, and tumor cells, M2d secrete proteases (such as MMP-2), cytokines (such as VEGF), and anti-inflammatory factors (such as TGF-β and IL-10) to promote angiogenesis and immunosuppression. In this study, we observed that reducing lactate secretion and transport decreased the intracellular lactate concentration in macrophages, resulting in a significant reduction in M2 polarization. This preliminary evidence supports the notion that PC cells promote M2 polarization by producing large amounts of lactate. Importantly, we discovered the M2 is characterized by high secretion of IL-10 and VEGF-A, which are the primary factors secreted by M2d. Our study confirmed that M2d induced by PC possess strong abilities to suppress CD8^+^ T cell function and promote microvascular formation. Compared to traditional therapies that simply inhibit M2 polarization, identifying specific targets to reverse M2d polarization may better address the issue of PC’s resistance to chemotherapy and immunotherapy.

METTL3 is a key component of the m^6^A methyltransferase complex, influencing cellular function and phenotype by regulating the m^6^A modification levels of different genes(16). Some studies suggest that METTL3 acts as an oncogene, promoting cell proliferation and invasion and increasing PC cell resistance to chemotherapy(32). Taketo et al. found that the loss of METTL3 enhances the sensitivity of PC cells to chemotherapeutic drugs such as gemcitabine, 5-fluorouracil, and cisplatin, as well as to radiotherapy(33). Recently, Jia Xiong et al. discovered that lactate in the TME of colon cancer not only promotes METTL3 expression through histone H3K18 lactylation but also lactylates the zinc finger domain of METTL3. This lactylation enhances the mRNA methylation of Jak1, which, through binding with YTHDF1, increases translation efficiency and activates the JAK1-STAT3 signaling pathway, creating an immunosuppressive environment. This reveals a novel mechanism of lactylation modification regulating the immunosuppressive function of tumor-infiltrating myeloid cells and mediating tumor immune escape(17). However, the regulation of METTL3 by lactate and the relationship between METTL3-mediated m^6^A methylation and the high infiltration of TAMs in PC remain unclear.

In our study, we found that co-culturing macrophages with PC cells significantly increased METTL3 expression in a lactate concentration-dependent manner. METTL3 expression was also elevated in regions of PC tissues with high M2 cell infiltration, showing a clear co-localization. When METTL3 was knocked down, the levels of PD-L1, IL-10, and VEGF-A in macrophages stimulated with CM were decreased. Correspondingly, the macrophages’ ability to inhibit CD8^+^ T cells and promote microvascular formation also diminished. Emerging studies have shown that TAMs interact with CD8^+^ T cells to reduce their mobility and limit their entry into tumors(34). In vivo, knocking down METTL3 or inhibiting it with STM2457 significantly slowed pancreatic cancer progression in mouse models, IF of mouse tumors revealed that METTL3 inhibition not only suppressed tumor growth but also reduced CD163+ M2 infiltration and increased CD8+ T cell infiltration. These results preliminarily confirm that PC cells may promote M2d polarization by upregulating METTL3 expression in macrophages.

Based on TAM’s signature genes, enriched pathways, and predicted functions, TAMs can be further categorized into seven subsets including interferon-primed TAMs (IFN-TAMs)(35). IFN-TAMs exhibit high expression of interferon-regulated genes and function as immunosuppressive macrophages, curbing immune responses through tryptophan degradation and the recruitment of immunosuppressive Tregs(36). Utilizing RNA-seq and MeRIP-seq technologies, we identified OAS3 as a pivotal target through which METTL3 modulates M2d polarization. OAS3, a critical immune mediator and a member of the enzyme class induced by interferons, may play a role in promoting IFN-TAM polarization and enhancing their pro-tumor functions in PC. Further investigation is required to elucidate whether OAS3 can facilitate IFN-TAM polarization and augment their tumor-promoting activities in pancreatic cancer. Li et al. found that OAS3 expression positively correlates with immune checkpoint-related genes in most tumors, suggesting that OAS3 may serve as a crucial biomarker for tumor immunotherapy. OAS3 is closely associated with pathways including the tumor inflammation signature, cellular response to hypoxia, tumor proliferation signature, angiogenesis, and apoptosis. Additionally, OAS3 expression correlates with the prognosis and chemotherapeutic outcomes of various cancers(19). Zhang et al. observed that M2 increase in dermatomyositis (DM) and myocarditis compared to healthy controls, associating with elevated OAS3 expression in these conditions(37). Numerous studies reporting a negative correlation between OAS3 and prognosis in various cancers, including PC, and its positive association with immunosuppressive cell infiltration, however, the regulatory mechanisms remain unclear(20). In this study, we found that the proportion of CD8^+^ T cells in lymphocytes significantly increased in the sh-OAS3 macrophage group, especially the TNF^+^GZMB^+^CD8^+^ T cell subset. Subsequently, we constructed an immune system humanized mouse CDX model and found that specific knockdown of OAS3 in macrophages synergized with Gem and PD-L1 mAb treatment. This is the first evidence confirming that PC cells promote M2d infiltration by upregulating OAS3 expression in macrophages. This discovery provides new theoretical support for targeting OAS3 as a potential therapeutic strategy.

However, this study has several limitations. Firstly, we could not completely exclude other factors that may also facilitate METTL3 upregulation, since the TME contains various effectors that educate myeloid cells to form an immunosuppressive network. Secondly, although our study demonstrated that OAS3 is regulated by METTL3, with OAS3 mRNA and protein expression decreasing upon METTL3 knockdown and MeRIP-seq revealing two OAS3 methylation sites significantly reduced after METTL3 knockdown, these methylation sites were not further validated in this study. The effects of METTL3-mediated m^6^A modification can vary depending on the reader proteins, leading to either increased mRNA stability or degradation. Whether METTL3 regulates OAS3 mRNA stability in macrophages through this mechanism and the specific reader proteins involved were not identified in our study. OAS3 is known to regulate apoptosis, and our study found that CM stimulation inhibits macrophage apoptosis, mainly reducing M2 macrophage apoptosis, which may be another potential mechanism for high M2 infiltration. Therefore, the detailed functional mechanisms of OAS3 in promoting M2d need further investigation. Finally, we observed that IL-6 secretion levels increased in the macrophage culture supernatant under CM stimulation. IL-6 is an important cytokine that promotes M2d polarization, and M2d may further enhance IL-6 release, forming a positive feedback loop. Both METTL3 and lactate inhibition can partially suppress IL-6 secretion, suggesting that this may be another potential mechanism by which lactate upregulates METTL3 to promote M2d polarization, warranting further exploration.

## Conclusions

Taken together, these studies establish a paradigm wherein PC cells derived lactate upregulate METTL3 in macrophages, thereby increasing the m^6^A methylation and expression levels of OAS3. Elevated OAS3 expression promotes the polarization of macrophages into CD163^high^M2d that secrete high levels of IL-10 and VEGF-A. This polarization inhibits TNF^+^GZMB^+^CD8^+^ T cells and promote neovascularization, ultimately facilitating PC progression (**Fig. 8**). Targeting OAS3 holds promise for reversing the polarization and infiltration of M2d in PC, remodeling the immunosuppressive TME, and ultimately improving the efficacy of clinical treatment.

## Methods

### Human samples

Human tissues from pancreatic cancer patient were received from Shanghai General Hospital performed in strict accordance with the recommendations of the protocol, which was approved by Institutional Review Board (No.2021SQ099).

### Cell lines

Human THP-1 monocytes, human PC cell lines (PANC-1、BXPC-3、AsPC-1), pancreatic ductal epithelial cells (Hpne), HUVECs and Panc02 cell line were purchased from Institute of Biochemistry and Cell Biology, Chinese Academy of Sciences (Shanghai, China). Specially, THP-1 cells were maintained in RPMI 1640 culture medium containing 10% FBS and 50pM β-mercaptoethanol, other cell lines were grown in DMEM media supplemented with 10% FBS and 1% penicillin and streptomycin in a 5% carbon dioxide cell incubator. For bone-marrow derived macrophages (BMDMs), mouse bone marrow was collected by flushing the femurs of 8-10 week old C57BL/6 mice with cold PBS. After collection, red blood cells (RBCs) were lysed with RBC lysis buffer (Beyotine,USA), and the remaining cells were washed twice with PBS.

### Induction of macrophage differentiation

Human THP-1 monocytes were differentiated into macrophages by treating with 100 ng/mL phorbol-12-myristate-13-acetate (PMA) (MCE, USA) for 24 hours. After differentiation, the Mφ polarized macrophages were cultured in fresh RPMI 1640 medium with 10% FBS for an additional 24 hours. For the induction of macrophage differentiation from bone marrow cells, the sorted cells were cultured in DMEM supplemented with 10% FBS and 20ng/mL mouse macrophage colony-stimulating factor (M-CSF) (Peprotech, USA). On day 6, the medium was switched to RPMI 1640 supplemented with 10% FBS, and the cells were treated with either DMSO or STM2457 for another 48 hours before harvesting.

### Macrophage polarization studies

In the co-culture experiments, THP-1 monocytes were differentiated into Mφ in 6-well Transwell inserts (membrane pore size of 0.4 μm, Corning, #3450) and then co-cultured with Hpne or human PC cells for 48 hours. For the preparation of tumor-conditioned media (TCM), PANC-1 cells were grown to 80% confluence, washed with PBS, and the medium was changed to DMEM without FBS for another 48 hours. The TCM was then collected and used for co-culture with Mφ induced from THP-1 cells. Finally, M2 (CD11b^+^CD163^+^) polarization levels were assessed, or the cells were collected for further analysis.

### Animals and *in vivo* experiments

C57BL/6, BALB/c, and NSG mice used for cells/drug treatment studies were obtained from the Experimental Animal Center of Shanghai General Hospital. One million Panc02 cells were subcutaneously injected into 6-8 week-old BALB/c or C57BL/6 mice. In the BALB/c mice group, Panc02 cells were mixed with either DMSO or STM2457 (25 μM) (MCE, HY-134836) pretreated BMDM (5×10^5^ cells/mouse) and then injected subcutaneously into the right axilla of the mice. C57BL/6 mice bearing tumors received intraperitoneal (i.p.) injections of clodronate liposomes (150 μl/animal, every 3 days) to deplete macrophages, either alone or in combination with STM2457 (dissolved in corn oil, 30 mg/kg, 3 times a week) to inhibit METTL3. NSG mice received i.p. injections of huPBMC (1×10^6^ cells/mouse), followed by the establishment of the PANC-1 CDX model. These mice were then treated with Mφ or sh-OAS3Mφ, either alone or in combination with Gem (20 mg/kg, 3 times a week) (MCE, HY-B0003) and anti-PD-L1 mAb (100 mg/kg, MCE, HY-155959).

### Flow cytometry

For flow cytometry staining, dissociated cells were incubated with Cell Staining Buffer (BioLegend, BD) followed by staining with primary antibodies directed against CD11b (Cell Signaling, 85601S), AnnexinV-FITC Apoptosis Detection Kit (Beyotime, C1062S) and CD163 (BD, 333606), CD86 (BD, 374208), PD-L1 (BD, 329706), CD8 (BD, 344705, 344732), anti-human CD45 (BD, 333606), anti-mouse CD45 (BD, 103130), CD3 (BD, 300317), CD4 (BD, 980802), Granzyme B (BD, 515403) and TNF-α (BD, 502943) were obtained from BD. For measurement of Granzyme B and TNF-α, cells were stimulated by Ionomycin (1μg/mL) (Yeasen, 50402ES03) and PMA (10ng/mL) (Yeasen, 50601ES03) followed by a 4−6h incubation at 37 ℃ according to the manufacturer’s instructions. Cells were then fixed and permeabilized in cytofix permeabilization solution and stained with Granzyme B and TNF-α antibody. Samples were analyzed by FACS after staining and data were analyzed using FlowJo software.

### Quantification and inhibition of Lactate

Lactate concentrations were determined using a colorimetric lactate assay kit according to the manufacturer’s protocol (Solarbio). To inhibit lactate production, PANC-1 cells were treated with 20mM 2-Deoxy-D-glucose (2-DG, HY-13966, MCE) for 24 hours. The medium was then replaced with fresh DMEM, and the lactate concentration in the TCM from PANC-1 cells was measured. During the culture of macrophages (Mφ) with TCM, we used the MCT1 selective inhibitor BAY-8002 (HY-122312, MCE) at a concentration of 1μM to inhibit lactate uptake by Mφ. After 48 hours, the intracellular lactate concentration in Mφ was measured.

### Immunofluorescence

Tissue sections of human PC were obtained from general department of Shanghai General Hospital. 5 mm formalin-fixed paraffin embedded tumor tissues sections and cells were blocked with blocking solution for 10 minutes. The human tissues were then stained with antibodies specific for CD163 (ab87099, 1:1200; Abcam) and METTL3 (ab217109, 1:1000; Abcam), the tumor tissues of C57BL/6 mice were stained with CD163 (ab87099, 1:1200; Abcam) and CD8 (ab217344, 1:2000; Abcam), the tumor tissues of NSG mice were stained with CD163 (ab87099, 1:1200; Abcam) and CD8 (85336, 1:1000; CST). For cells, they were incubated with CD163 (ab87099, 1:1200; Abcam) and METTL3 (ab217109, 1:1000; Abcam) antibodies for 1 hour at room temperature. After 3 washings with TBST buffer, samples were stained with HRP polymer-conjugated secondary antibodies for 60 minutes at room temperature and then incubated with Alexa Fluor 488 and Alexa Fluor 594 reagents (Abcam) for 60 minutes. DAPI was used to counterstain the nuclei. The images of fluorescent staining were captured using Leica fluorescent microscope.

### Western blot analysis

Cells were collected and lysed, Cell lysates were separated on 10% SDS-PAGE gels and blotted on protran nitrocellulose membranes. Membranes were blocked with 5% milk in TBS-Tween and incubated overnight with the following primary antibodies: Anti-METTL3 ab (1:1000, ab195352, Abcam), OAS3 Polyclonal ab (1:1000, 21915-1-AP, Proteintech) and GAPDH ab (1:1000, ab181602, Abcam) followed by a 1 hour incubation with a secondary HRP-conjugated antibody and chemiluminescent detection using ECL kit (7E730K3, Vazyme). Quantification was performed using ImageJ.

### Quantitative real-time PCR

Total RNA was isolated from Mφ cells using the Trizol reagent (Life technology, USA) according to standard protocols, and then applied the Thermo Scientific K1622 Revert Aid First Strand cDNA Synthesis Kit. qRT-PCR was performed using the ChamQTM Universal SYBR qPCR Master Mix (Vazyme Bioteh Co.,Ltd) according to standard protocols. All primers used in this study were acquired from AZENTA (Suzhou, China), relative expression levels were normalized to Gapdh expression.

### RNA-seq

sh-NC and sh-METTL3 Mφ cells were generated by PMA as described above and co-cultured with TCM or 2-DG pretreated TCM. Total RNA was extracted using Trizol reagent kit according to the manufacturer’s protocol. The enriched mRNA was fragmented into short fragments and reversely transcribed into cDNA by using NEBNext Ultra RNA Library Prep Kit for Illumina (NEB #7530,New England Biolabs, Ipswich, MA, USA). The purified double-stranded cDNA fragments were end repaired, A base added, and ligated to Illumina sequencing adapters.The ligation reaction was purified with the AMPure XP Beads(1.0X). And polymerase chain reaction (PCR) amplified. The resulting cDNA library was sequenced using Illumina Novaseq6000 by Gene Denovo Biotechnology Co. DO enrichment analysis identified significantly enriched human disease DO terms in DEGs comparing with the whole genome background. All DEGs were mapped to GO terms in the Gene Ontology database (http://www.geneontology.org/), gene numbers were calculated for every term, significantly enriched GO terms in DEGs comparing to the genome background were defined by hypergeometric test.

### MeRIP-seq

After total RNA was extracted from sh-NC and sh-METTL3 Mφ cells, the enriched RNA was fragmented into short fragments (about 100nt) by using fragmentation buffer. RNA was divided into two parts, one of which was used as the input (no IP experiment was performed). The other RNA was enriched with m^6^A specific antibody (EpiMark N6-Methyladenosine Enrichment Kit, E161OS, NEB). The enriched RNA was reverse transcripted into cDNA with random primers. Next, the cDNA fragments was end repaired, and ligated to Illumina sequencing adapters. Finally we got qualified library for sequencing, sequenced using Illumina NovaSeqTM 6000 (or other platforms) by Gene Denovo Biotechnology Co. The interaction between protein and RNA were not random, while they showed some specific sequence preference. MEME suit (http://meme-suite.org/) and DREME (http://memesuite.org/tools/dreme) were used to detect the significant sequence motif in the transcript sequence associated with peaks. The exomePeak2 software was used to analysis RNA methylation rate between groups. The relative methylation rate of each peak was calculated using MeRIP data and Input data. According to the genomic location information and gene annotation information of peak, peak related genes can be confirmed. Besides, the distribution of peak on different function regions, such as 5’UTR, CDS and 3’UTR, was performed.

### Source and preprocess of publicly attainable PC expression datasets

The publicly available NCBI Gene-Expression Omnibus (GEO) datasets (https://www.ncbi.nlm.nih.gov/geo/) and the Cancer Genome Atlas (TCGA) (https://cancergenome.nih.gov/) were utilized to retrospectively gather gene expression data and clinical characteristics of PC samples. Samples lacking survival information were excluded from further evaluation. Three eligible PC cohorts (GSE28735, GSE62452, TCGA) were included in this study for comprehensive analysis(38,39). For the TCGA datasets, RNA sequencing data (FPKM values) were downloaded using the TCGAbiolinks package, designed specifically for integrating GDC data. FPKM values were converted to transcripts per kilobase million (TPM) values. To mitigate batch effects across different datasets, the “Combat” algorithm from the R package sva was employed(40).

### Functional annotation and gene set variation analysis (GSVA)

GSVA enrichment analysis was conducted using the “GSVA” package in R to explore the heterogeneity within the OAS3 high and low expression groups(41). GSVA, a non-parametric and unsupervised method, is widely utilized to assess variation in pathway activities and biological processes across expression datasets. This analysis involved gene sets “c2.cp.kegg.v7.5.1.symbols” from MSigDB, using the GSVA package version 1.42.0.

### Analysis of TME immune cell infiltration

The ssGSEA (single sample gene set enrichment analysis) algorithm was utilized to assess the relative abundance of immune cell infiltration within the TME and to evaluate immune function in PC patients(42). Additionally, the ESTIMATE algorithm was employed to estimate the presence of immune and stromal cells in PC(43). This algorithm facilitates the prediction of immune and stromal cell infiltration levels by calculating respective immune and stromal scores.

### Single-cell RNA sequencing (scRNA-seq) analysis

The dataset CRA001160 was downloaded and initially processed using the “Seurat” R package for filtering and standardization(44). Post-standardization, we retained the 1500 genes with the highest variance for further analysis. Dimensionality reduction was performed via PCA, followed by cell clustering using UMAP. Cell annotations were assigned using the “SingleR” R package, referencing CellMarker(45). Single-cell pseudotime trajectories were constructed with Monocle 2 (2.10.1)

### Lentiviral transfection

Lentiviral vectors encoding shorthairpin RNAs (shRNAs) against METTL3 (sh-METTL3) and OAS3 (sh-OAS3) were obtained from Shanghai Jiaotong University School of Medicine (Shanghai, China). The stably transfected cell lines were selected by adding 2 ug/ml of puromycin (Beyotime, Guangzhou, China) into the cells for 2 days.

### ELISA assays

Culture supernatants from PANC-1 cells were assayed for total IL-10,VEGF-A, IL-6, IL-1b and IFN-γ by using commercially available Legend Max ELISA Kits from BioLegend according to manufacturer’s instructions.

### Tube formation assays

A Matrigel tube formation assay was performed as previously described(46). In brief, serum-free DMEM and growth factor-reduced basement membrane matrix (BD Biosciences) were mixed in a 1:1 ratio and seeded into 96-well plates, followed by incubation at 37℃ for 30 minutes to allow gel formation. HUVECs were then harvested and resuspended with the conditioned media from different treatment groups of macrophages. The cell suspension was seeded into the prepared 96-well plates at a density of 20,000 cells per well. The plate was further incubated at 37℃ for 4 hours, and the tube formation was visualized under an inverted microscope. Enclosed networks of tube structures from three randomly chosen fields were recorded under a light microscope.

### Statistical analysis

For all *in vitro* studies, experiments were performed three or more times with three or more biological replicates per group. All *in vivo* experiments were performed with 4 to 5 animals assigned/group. Data were normalized to standard where applicable. Significance testing was performed by one-way ANOVA with Tukey post hoctestingfor multiple pair-wise testingwith more than two groups and by Student t test when only two groups were compared. Statistical analysis was performed using GraphPad.Prism.8 software. Unless otherwise stated, data are shown as mean SEM. In all cases, *, *P* < 0.05, **, *P* < 0.01, ***, *P* < 0.001, and ns, no significance.

## Ethics approval and consent to participate

This study was approved by the Institutional Review Board of Shanghai General Hospital. Written informed consent was obtained from all legal guardians of the patients. All procedures involving animals were approved by the Animal Welfare & Ethics Committee of Shanghai General Hospital (No. 2023AW024).

## Data availability

All data generated or analyzed during this study are included in this published article and its supplementary information fles.

## Acknowledgements

This work was supported by grants from the National Natural Science Foundation of China.(Grant No. 82173163, 81974372, and 82203751). We appreciate Long Jiang and Zheng Yan who provide clinical samples and help for this research.

## Author contributions

Shaopeng Zhang: Data curation, Formal analysis, Investigation, Methodology, Software and Writing - original draft. Ximo Xu: Formal analysis, Investigation, Methodology and Writing - original draft. Kundong Zhang: Formal analysis, Investigation, Methodology and Writing - original draft. Changzheng Lei: Investigation, Methodology and Software. Yitian Xu: Investigation, Methodology and Software. Pengshan Zhang: Funding acquisition, Methodology, Writing - review & editing. Yuan Zhang: Methodology, Writing - review & editing. Haitao Gu: Formal analysis, Resources, Supervision, Validation and Writing - review & editing.Chen Huang: Conceptualization, Data curation, Formal analysis, Project administration, Supervision and Writing - review & editing. Zhengjun Qiu: Conceptualization, Formal analysis, Funding acquisition, Investigation, Project administration, Resources, Supervision and Writing - review & editing.

---

**Supplementary Figure 1.**
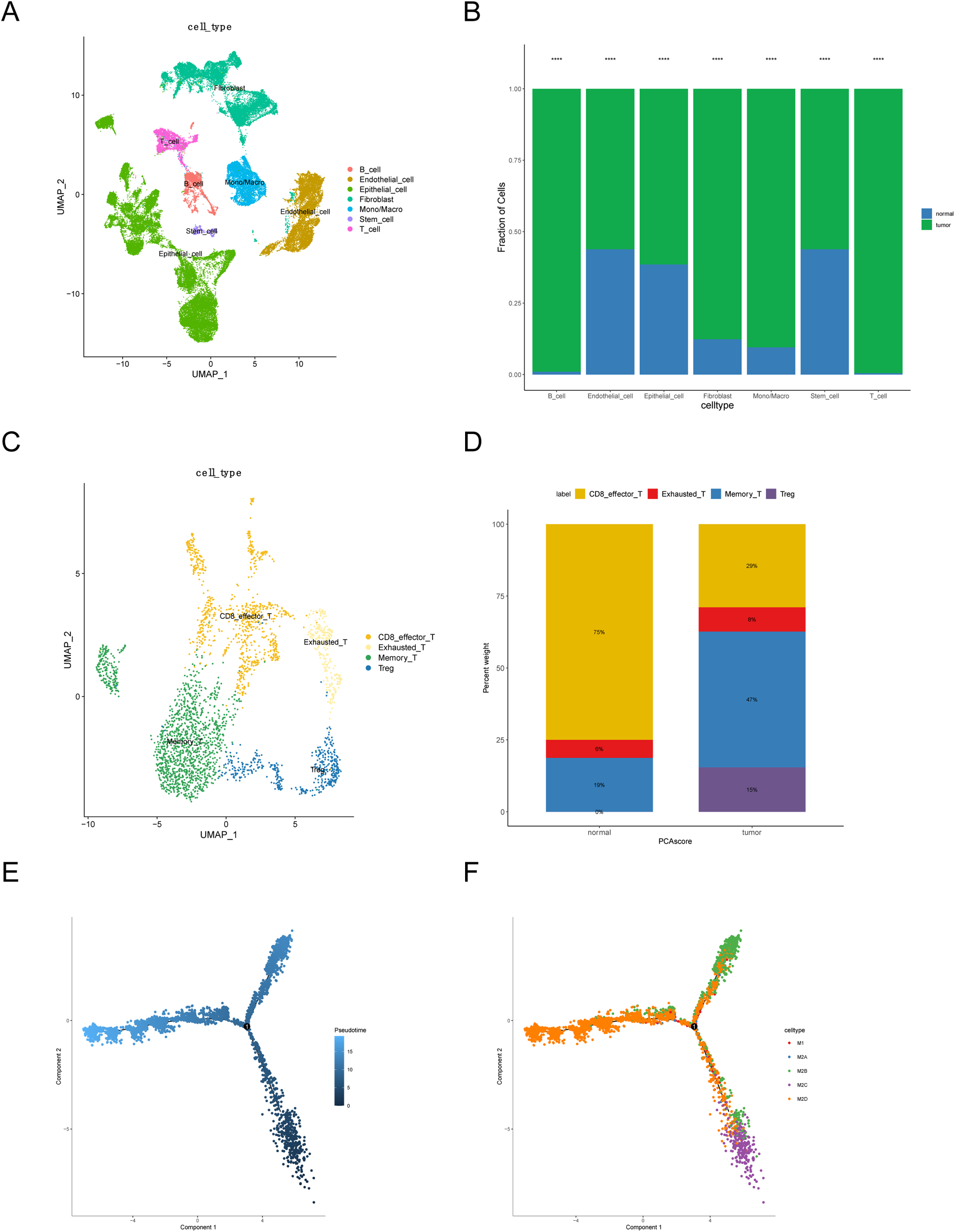
A. The UMAP plot of cells grouped into 7 phenotypes. B. Percentage of each distinct cells in the tumor and normal PDAC samples. C. The UMAP plot of T cells grouped into 4 phenotypes. D. Percentage of each distinct T cells in the tumor and normal PDAC samples. E-F. Monocle 2 trajectory analysis of macrophages obtained by 5’ gene expression chemistry annotated by calculated states.

**Supplementary Figure 2.**
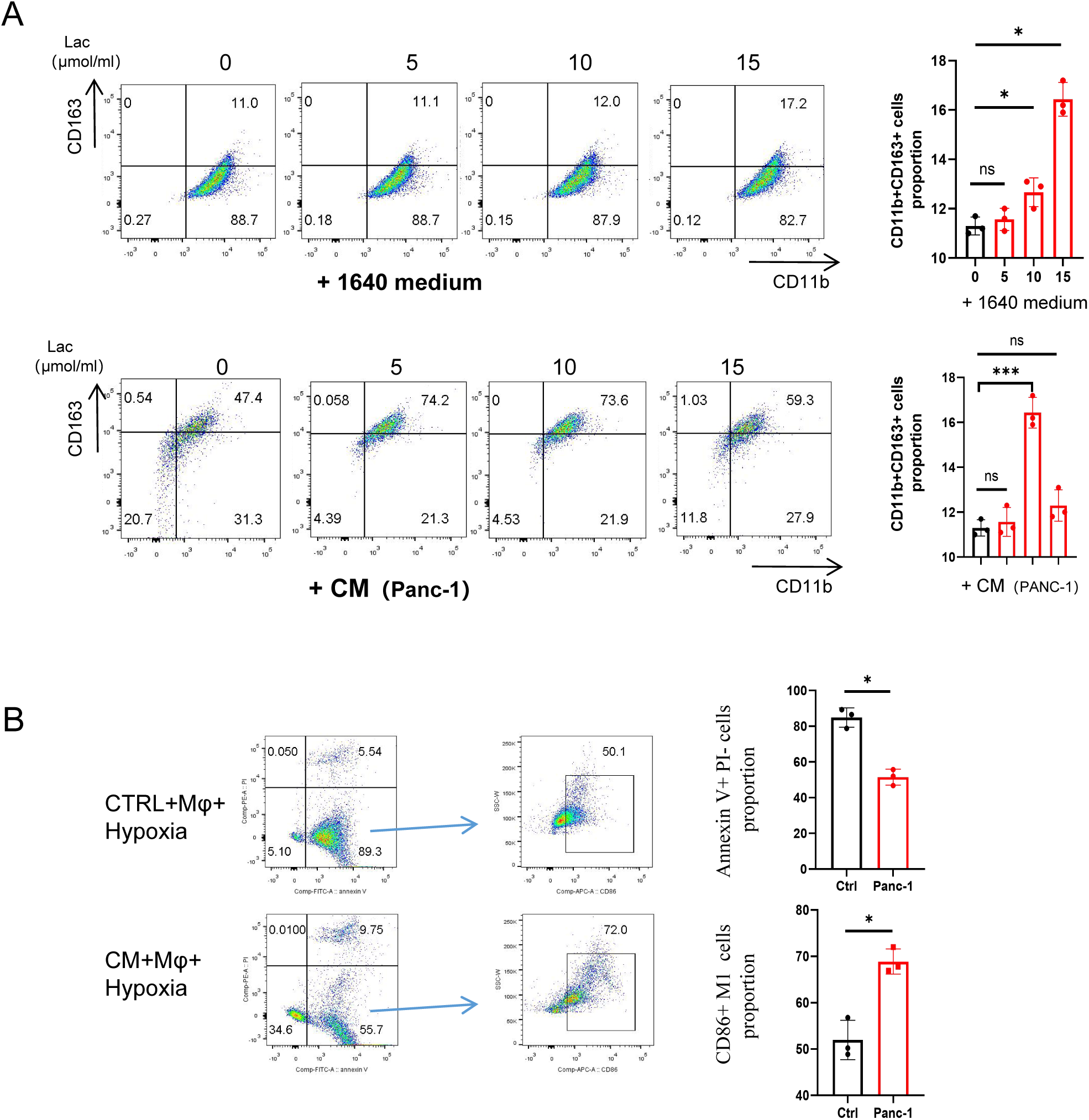
Left: Flow cytometry analysis showing the differences in M2 polarization levels under varying lactate concentrations in macrophage culture systems with regular 1640 medium and 1640 medium conditioned with PANC-1 cells (CM). Right: Proportion of CD11b^+^CD163^+^ M2 in different groups (n=3). B. Comparison of apoptosis levels in macrophages under hypoxic conditions with normal 1640 medium and PANC-1 cell-conditioned medium (CM). Statistical significance was determined by Student t test. *, *P* < 0.05, ***, *P* < 0.001, ns., no significance.

**Supplementary Figure 3.**
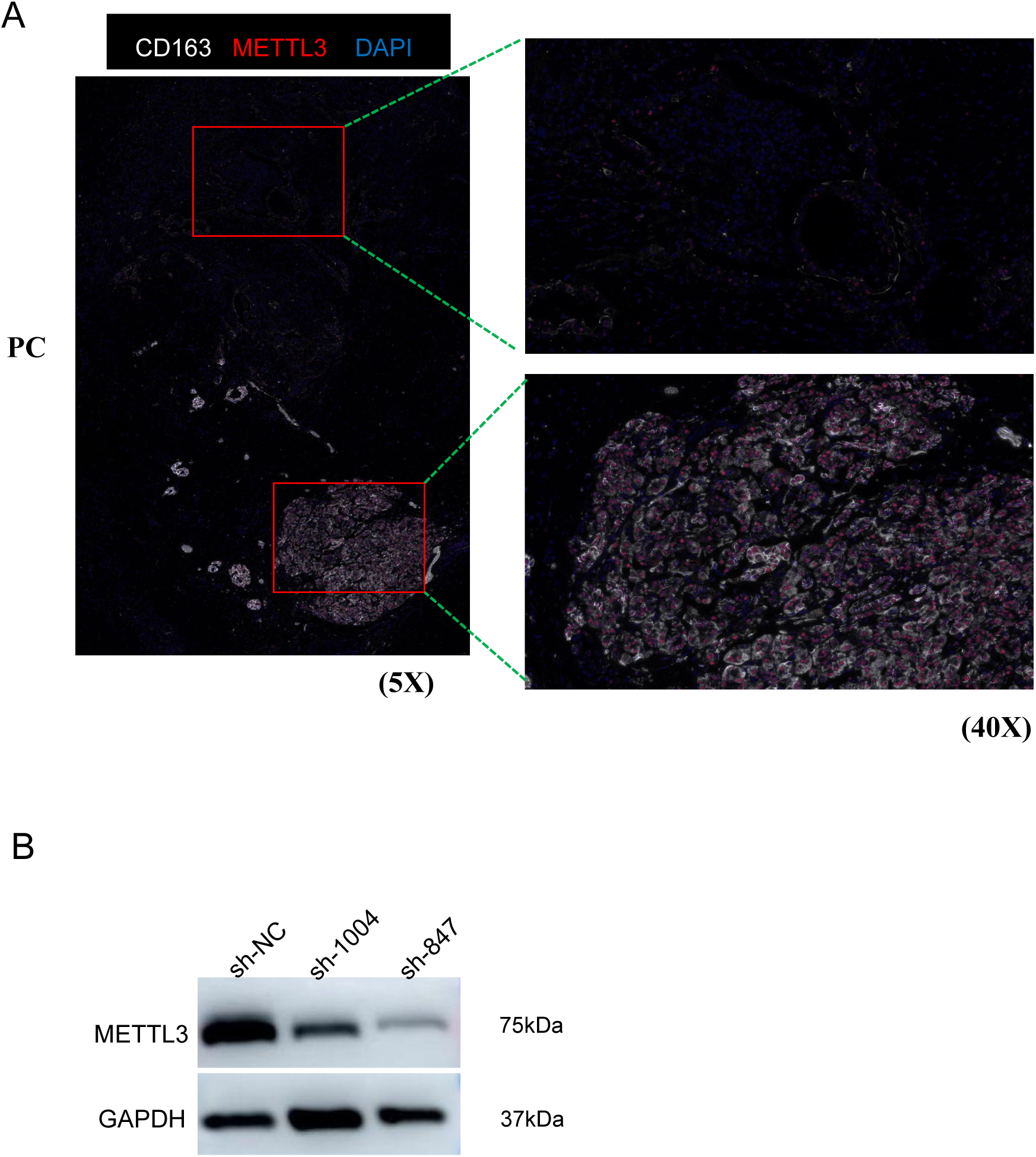
IF staining colocalization of CD163 and METTL3 in a human PDAC patient sample. B. Western blot images showing the expression of METTL3 in Mφ after treatment with lentiviral vectors encoding shorthairpin RNAs (shRNAs) against METTL3 (targeting sites 1004 and 847).

**Supplementary Figure 4.**
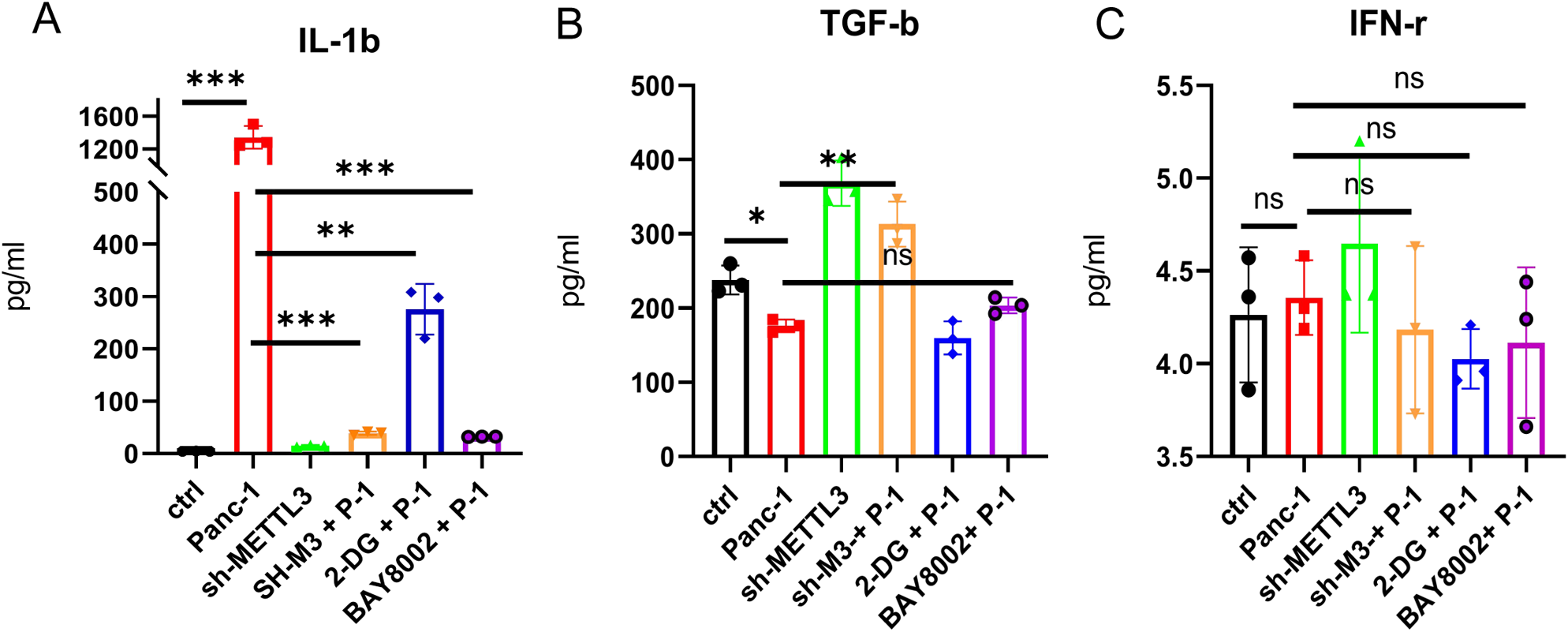
A-C. ELISA measurements of IL-1β (A), TGF-β (B), and IFN-γ (C) release under different culture conditions in macrophages and sh-METTL3 macrophages (n=3). Statistical significance was determined by Student t test. *, *P* < 0.05; **, *P* < 0.01, ***, *P* < 0.001, ns., no significance.

**Supplementary Figure 5.**
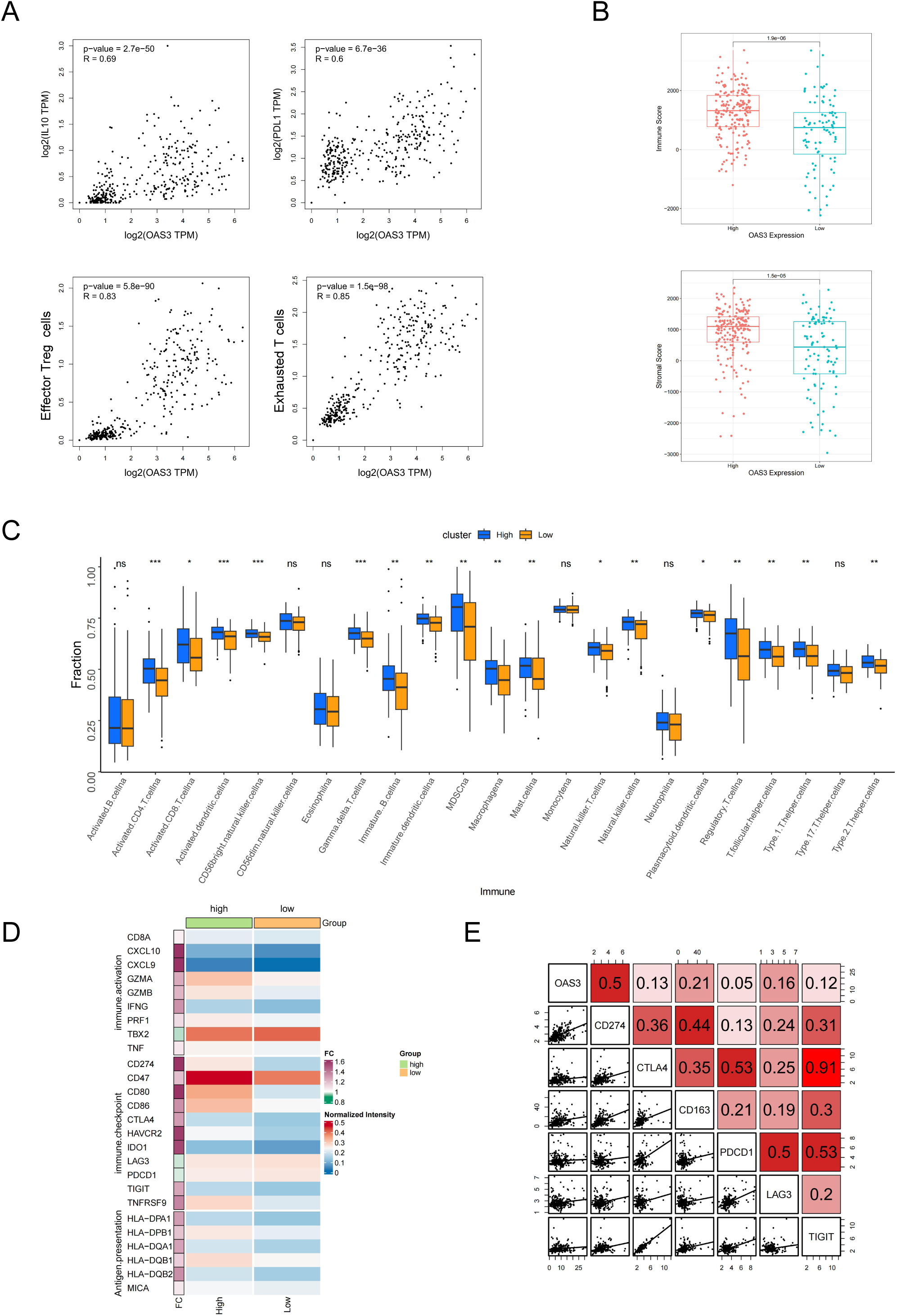
A. Scatter plots showing positive correlations between OAS3 expression and IL-10, PD-L1, Treg cells, and exhausted T cells. B. The immune score and stromal score of OAS3 high and low expression groups using Estimate algorithm in TCGA-PDAC cohort. C. The fraction of TME cell infiltration of OAS3 high and low expression groups using the ssGSEA algorithm. The top end, median line, and bottom end of the box represent the 25%, 50%, and 75% value, respectively. D. Correlation analysis between OAS3 expression levels and T cell exhaustion related markers expression levels. E. The comparison of expression level of antigen presentation, immune-activation and immune-checkpoint gene sets between OAS3 high and low expression groups. The asterisks illustrate the statistical p-value (*, *p* < 0.05, **, *p* < 0.01, ***, *p* < 0.001, ns., no significance.).

**Supplementary Figure 6.**
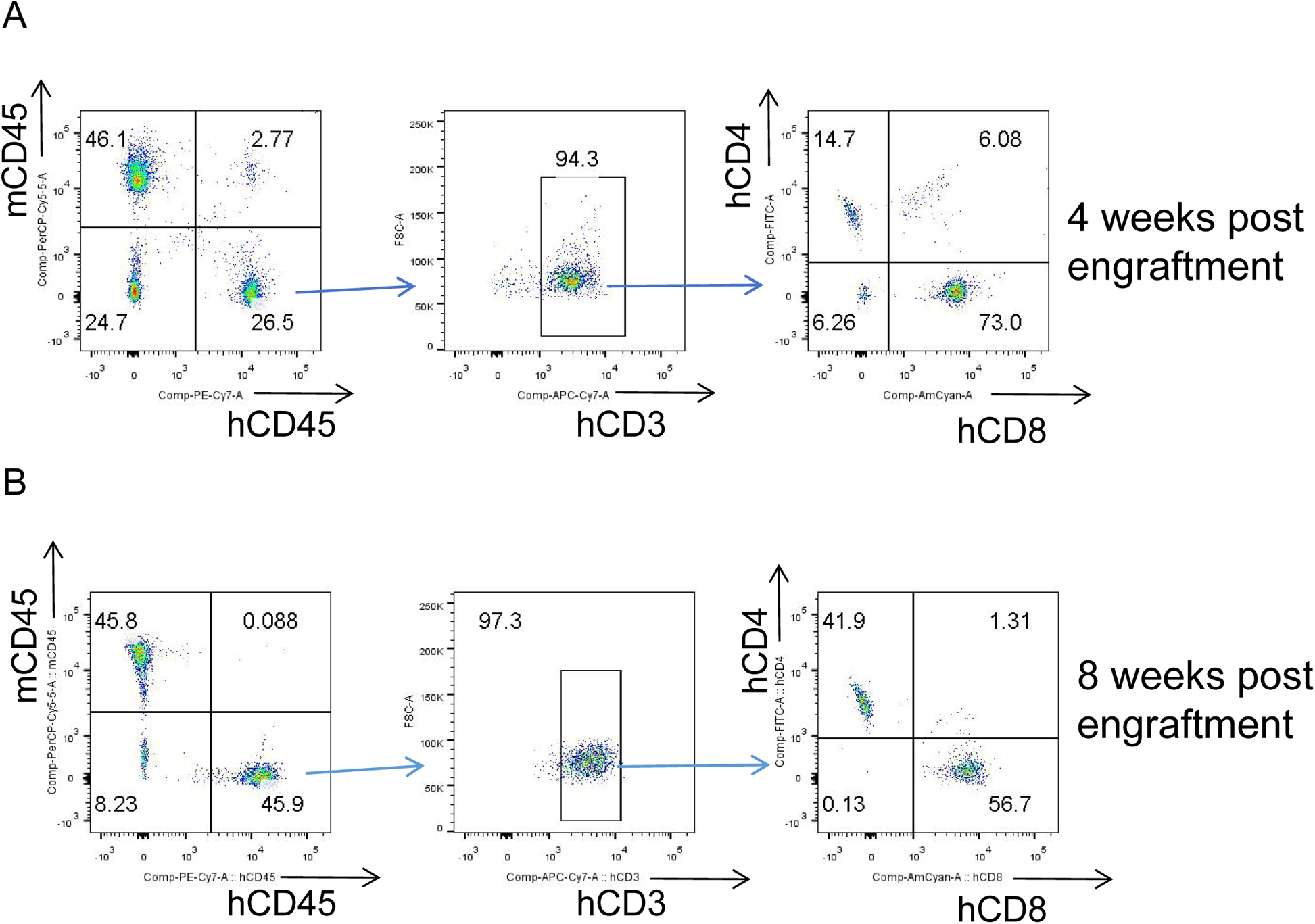
Flow cytometry analysis to detect the proportion of human CD3^+^CD4^+^ T cells and CD3^+^CD8^+^ T cells in peripheral blood of mice after intraperitoneal injection of PBMCs for 4 weeks (A) and 8 weeks (B).

